# Lifetime experiences as proxies of cognitive reserve predict cognition and motor function beyond multimodal MRI brain measures in healthy adults

**DOI:** 10.1101/2025.03.19.25324277

**Authors:** Isaac Saywell, Sabrina Sghirripa, Angela Walls, Andrew Dwyer, Brittany Child, John Salamon, Lyndsey Collins-Praino, Mark Jenkinson, Irina Baetu

## Abstract

Individual differences in lifetime experiences may moderate the association between brain characteristics and task performance, although the interplay between these factors and behavioural outcomes is unclear. Using multimodal magnetic resonance imaging (MRI), we explored whether lifetime experiences, as a cognitive reserve (CR) proxy, or brain measures more strongly predict cognition and motor function in healthy adults. A total of 101 subjects completed lifetime experiences, cognition, and motor function assessments. Structural, diffusion, and functional imaging measures were investigated. Associations between lifetime experiences, brain measures, and outcomes were explored using multiple linear regression. Controlling for demographics, CR proxy score was a significant moderator of the relationship between structurally derived brain measures and both cognition and motor function. CR proxy score was the only significant predictor of cognition and motor function, explaining variance beyond brain characteristics. MRI brain measures were not associated with overall cognition, overall motor function, or CR proxy score. CR proxies accounted for more variance in global cognition and all motor tests compared to brain measures; however, specific cognitive outcome test performance variance was more nuanced, often more strongly associated with brain measures. These findings highlight, for the first time, that enriching lifetime experiences account for differences in cognitive and motor function among healthy adults over and above what structural and functional brain characteristics can explain. Thus, CR appears to not only protect against brain changes, but also provides functional advantages in the absence of pathology, suggesting that greater engagement in stimulating experiences enhances healthy adult quality of life.

## 1 Introduction

Ageing is an incredibly heterogenous process. In older populations, functional abilities vary considerably compared to younger adults (Ferrucci & Kuchel, 2021), and individual differences in outcomes have been well-documented in longitudinal studies (Cai et al., 2022; Schaie & Willis, 2010; Wilson et al., 2002). These individual differences in performance have also been observed in neurodegenerative conditions, such as Alzheimer’s disease and Parkinson’s disease (Nelson et al., 2021; Saywell et al., 2024), where a type of reserve is thought to sustain task performance, despite the accumulation of brain pathology (Katzman et al., 1988). Two main models of reserve have been proposed. Brain reserve acts passively, suggesting that neurobiological capital helps resist brain damage until a fixed threshold is reached (Stern, 2002). Reserve has also been suggested to function actively in response to pathology, a concept termed cognitive reserve (CR). According to this model, CR grants more efficient usage of cognitive and functional brain processes, thereby increasing neural capacity, which is linked to better task performance and enhanced ability to cope with pathology (Stern, 2009).

Directly measuring the mechanisms underlying CR is challenging due to its theoretical nature. Instead, CR is usually quantified indirectly through lifetime experiences, such as education, occupation, leisure time activities, social engagement, and verbal intelligence (Stern, 2002), which are thought to underpin CR by giving rise to neuroprotective cognitive strategies (Jones et al., 2011; Valenzuela & Sachdev, 2006b, 2006a). These factors are widely used as proxy measures of CR, as they have demonstrated longitudinal associations with preserved cognitive performance in normal and pathological ageing. The most commonly reported CR proxy, years of education, moderates the relationship between pathology and cognition in older adults, such that those with higher educational attainment are better able to maintain cognitive function despite age-associated brain changes (Querbes et al., 2009; Rentz et al., 2010). Further, given that performance in motor tasks typically requires cognitive planning (Kobayashi-Cuya et al., 2018; Leisman et al., 2016), lifetime experiences may influence motor function in healthy adults. In older adults, those with more years of education perform better in motor function assessments (Elbaz et al., 2013), and higher CR composite measure scores are correlated with faster responses in motor tasks (Quinzi et al., 2020). However, interpretations based solely on simple correlations are limited, as a proxy’s effect could reflect factors unrelated to CR (Jones et al., 2011). Sufficiently evaluating CR requires either examining the interaction between a proxy and brain status, or demonstrating that a proxy accounts for outcome variance beyond brain characteristics (Stern et al., 2023). Studies often investigate the relationship between CR proxies and behavioural outcomes without adequately considering neuroanatomical features that may drive this association. Consequently, it is difficult to determine whether individual differences in cognitive or motor function reflect the relative contributions of lifetime experiences or neurobiological factors.

Brain characteristics have been linked to individual differences in functional performance. Co-occurrence of age-related cognitive decline and neuroanatomical deterioration, in the form of atrophy, increased white matter (WM) diffusivity, and reductions in brain activity at rest, have been widely reported (Damoiseaux et al., 2008; Madden et al., 2012; Nyberg et al., 2012; Salthouse, 2011). Age-related alterations in motor function are also linked to the functional status of neural circuits, as well as cortical atrophy and WM tract integrity (for a review see Seidler et al. 2010). Furthermore, proxies of CR may be associated with brain characteristics. Of note, healthy adults with higher CR proxy composite scores have been shown to possess increased grey matter (GM) volume in the supplementary motor area, middle frontal gyrus, angular gyrus, rectus gyrus, middle cingulum and the cerebellum (Conti et al., 2021). These individuals also demonstrated decreased resting-state functional MRI (rsfMRI) connectivity (FC) in the sensorimotor, salience, and executive-control networks, but increased FC in the default mode network, indicating that lifetime experiences may differentially affect brain networks (Conti et al., 2021). In older adults, more years of education, greater occupational complexity, higher verbal intelligence, and increased engagement in stimulating leisure activities may be associated with greater tolerance to age-related changes in WM tract microstructure (Arenaza-Urquijo et al., 2011; Haut et al., 2007). Interestingly, in a longitudinal study, lifetime experiences were positively associated with WM tract integrity in younger individuals, but negatively associated in older adults when controlling for cognitive performance (Brichko et al., 2022), suggesting that the function of CR on microstructural changes may shift across the lifespan.

Despite established relationships between behavioural outcomes and both CR proxies and brain characteristics, the interplay among these factors remains poorly understood. Given the overlap between the associations that lifetime experiences and *in vivo* neuroanatomical measures have with task performance, it is important to establish whether these factors contribute to shared or exclusive variance. Further, it is estimated that targeting 14 modifiable lifestyle factors across the lifespan could prevent or delay the onset of approximately 40% of dementia cases worldwide (Livingston et al., 2024). Lifetime experiences that contribute to CR are thought to support brain processes that enhance performance in healthy younger adults and, later in life, help compensate for age-related brain changes (Tucker & Stern, 2011). Thus, understanding how stimulating experiences independently affect and shape behavioural outcomes, beyond brain characteristics, could inform interventions that optimise and preserve performance throughout the lifespan, therefore improving quality of life. Investigating CR proxies and neuroimaging-derived brain indices simultaneously would help elucidate whether discrepancies in outcomes can better be attributed to neuroanatomical variation or differences in lifetime experiences. Despite this, to the best of our knowledge, no previous study has used a multimodal magnetic resonance imaging (MRI) approach to explore if CR proxies contribute beyond brain measures to individual differences in cognition and motor function. Therefore, in the present study, which is the first of its kind, we aimed to identify neuroanatomical features, measured by multimodal MRI, associated with lifetime experiences, and whether the predictive utility of these lifestyle factors and brain measures varies across both cognition and motor function in healthy adults. We hypothesised that lifetime experiences are positively associated with brain characteristics, and that these CR proxies account for unique variance in behavioural outcomes beyond that explained by brain measures.

## 2 Methods

### 2.1 Subjects and data collection

A total of 101 healthy adults (44 males and 57 females) aged between 18 and 80 years (average age = 53.39 ± 16.98 years) were recruited between October 2020 and November 2024 from an ongoing cross-sectional study examining behavioural outcomes in normal ageing among 1150 individuals. The selected subset of study participants were those from the larger sample who underwent both an MRI scan and behavioural testing within six months of the scan. All subjects completed an online questionnaire prior to cognitive and motor function assessments. English-speaking individuals aged 18 or older, without a diagnosed neurological impairment and with no prior history of brain injury, were eligible. Other exclusion criteria included an uncorrected visual or hearing impairment, a diagnosed learning disability, history of drug or alcohol abuse, smoking more than five cigarettes per day, and, for the smaller subset who underwent an MRI scan, being ineligible to undergo an MRI scan (i.e., claustrophobic, pregnant, or possessing a metal implant). In rare cases where participants were missing data for a specific behavioural test, the median value of that test was imputed (see Supplementary Material Table 1 for an imputation count summary). Ethics approval was obtained from the University of Adelaide Human Research Ethics Committee (approval numbers H-2021-120 and H-2020-017), research was carried out according to the National Statement on Ethical Conduct in Human Research, and all participants provided signed informed consent.

### 2.2 Behavioural outcome measurements

#### 2.2.1 Cognition

A comprehensive battery of neuropsychological assessments was administered on an iPad. Working memory was tested visually and verbally through a dot matrix task (Law et al., 1995) and the digit span task (Woods et al., 2011), respectively. Visual processing speed was measured via inspection time (Vickers et al., 1972) and a two-choice reaction time task (Ratcliff & Smith, 2004). The two-choice reaction time task requires subjects to hold their dominant index finger on a blue circle before reacting to one of two squares by touching the one that lights up white. From this, decision time, the period between one square lighting up white and the participant lifting their finger, was derived as another measure of visual processing speed. Response time, a measure of motor speed (see section 2.2.2), was also determined using the two-choice reaction time task by calculating the duration between a subject lifting their finger off the blue circle and placing it on the square that changed colour to white. Raven’s matrices were used to estimate visuo-spatial reasoning ability (Raven & Raven, 2003). Stop signal reaction time (SSRT) was calculated using a stop signal task paradigm, consisting of ‘Go’ and ‘NoGo’ tasks, to measure response inhibition (Logan & Cowan, 1984). Time difference between incongruent and congruent trials on the Simon Task was used to assess executive functioning and selective attention (Lu & Proctor, 1995). Additionally, a researcher administered the Montreal Cognitive Assessment (MoCA; Nasreddine et al., 2005) as an evaluation of global cognitive ability. Higher scores on all cognitive tests indicated better performance. To accommodate for this, performance on speed-based assessments, such as the choice-reaction time task (decision time), inspection time, the Simon task, and the stop signal task, were inverted by subtracting each subject’s score from the maximum score in the sample.

#### 2.2.2 Motor function

Motor function was assessed across four different abilities: speed of hand movement, tremor, balance, and response time. A tapping test on an iPad was used to evaluate hand dexterity by capturing how fast a subject could move their index finger to alternate between tapping two static dots (3.5cm diameter) 13cm apart within 30 seconds. Average number of taps within the last 25 seconds of each 30-second interval (right and left hands) was computed as the final score. Tremor and balance were measured using a Nintendo® Wii remote and balance board, respectively, and custom recording software was used to process and store data. For tremor, participants were required to sit still and hold the controller limply, with their palm facing upward, for 60 seconds in each hand. Average theta power (3-7Hz) from both hands was used to estimate hand movement using the controller accelerometer, an accurate measurement of resting tremor (Daneault et al., 2013; Koçer & Oktay, 2016). For balance, subjects were asked to stand as stationary as possible on the board for 30 seconds with their feet together, while staring at a red dot positioned two metres away and 1.75 metres high. Path length, the total distance travelled by the centre of pressure, was used to generate a balance score, with a greater path length corresponding to worse balance. The Wii balance board has been validated against laboratory-grade force platforms (Clark et al., 2010), an approach which we also verified with our own paradigm (see Supplementary Material Fig 1). Balance data was not obtained for one subject due to a technical failure. Log transformation was applied to tremor and balance measures to account for any extreme outliers. Like cognition, higher scores on all motor function tests indicated better performance. Thus, assessments where higher raw scores are associated with worse motor function, like balance (path length), tremor (theta power), and the choice-reaction time task (response time), were inverted by subtracting each subject’s score from the maximum score in the sample.

### 2.3 MRI acquisition

MRI images were collected using either a Siemens Magnetom Skyra 3.0 T MRI scanner or a Siemens Magnetom Cima.X 3.0 T MRI scanner, both with a 64-channel head/neck coil, across a 45 minute brain scan protocol consisting of sequences: (1) 3D T1-weighted magnetisation-prepared rapid gradient echo sequence (5 minutes and 30 seconds scan time, 192 contiguous 1mm sagittal slices, voxel size = 1 x 1 x 1mm, echo time [TE] = 2.96ms, repetition time [TR] = 2300ms, inversion time = 900ms, flip angle = 9°, and field of view [FOV] = 256 x 256 x 192mm); (2) Diffusion-weighted images collected using standard echo-planar imaging with simultaneous multi-slice (6 minutes and 32 seconds scan time, 72 contiguous 2mm axial slices, voxel size = 2.02 x 2.02 x 2mm, TE = 94ms, TR = 3600ms, flip angle = 90°, FOV = 210 x 210 x 144mm) or CMRR multiband echo-planar imaging (6 minutes and 32 seconds scan time, 72 contiguous 2mm axial slices, voxel size = 2.02 x 2.02 x 2mm, TE = 94ms, TR = 3600ms, flip angle = 78°, FOV = 210 x 210 x 144mm) sequence, with diffusion gradients applied along 104 noncollinear directions using three b-values: 0 [B0 image], 1000s/mm2 and 2000s/mm2 in both sequences; (3) rsfMRI echo-planar imaging sequence using blood oxygen level dependent contrast with 490 volumes (6 minutes and 10 seconds scan time, 64 contiguous 2.4mm axial slices, voxel size = 2.4 x 2.4 x 2.4mm, TE = 39ms, TR = 736ms, flip angle = 52°, FOV = 210 x 210 x 154mm). Participants were instructed to keep their eyes open for the rsfMRI sequence.

### 2.4 MRI pre-processing

Neuroimaging data for each scan type was pre-processed using FMRIB Software Library (FSL; www.fmrib.ox.ac.uk) and FastSurfer (Henschel et al., 2020) tools. To eliminate bias, and account for contributions of many different brain characteristics, we opted for automated segmentation, atlas, and data-driven approaches. The final list of neuroanatomical features used in the analyses are presented in Fig 1.

**Fig 1.**
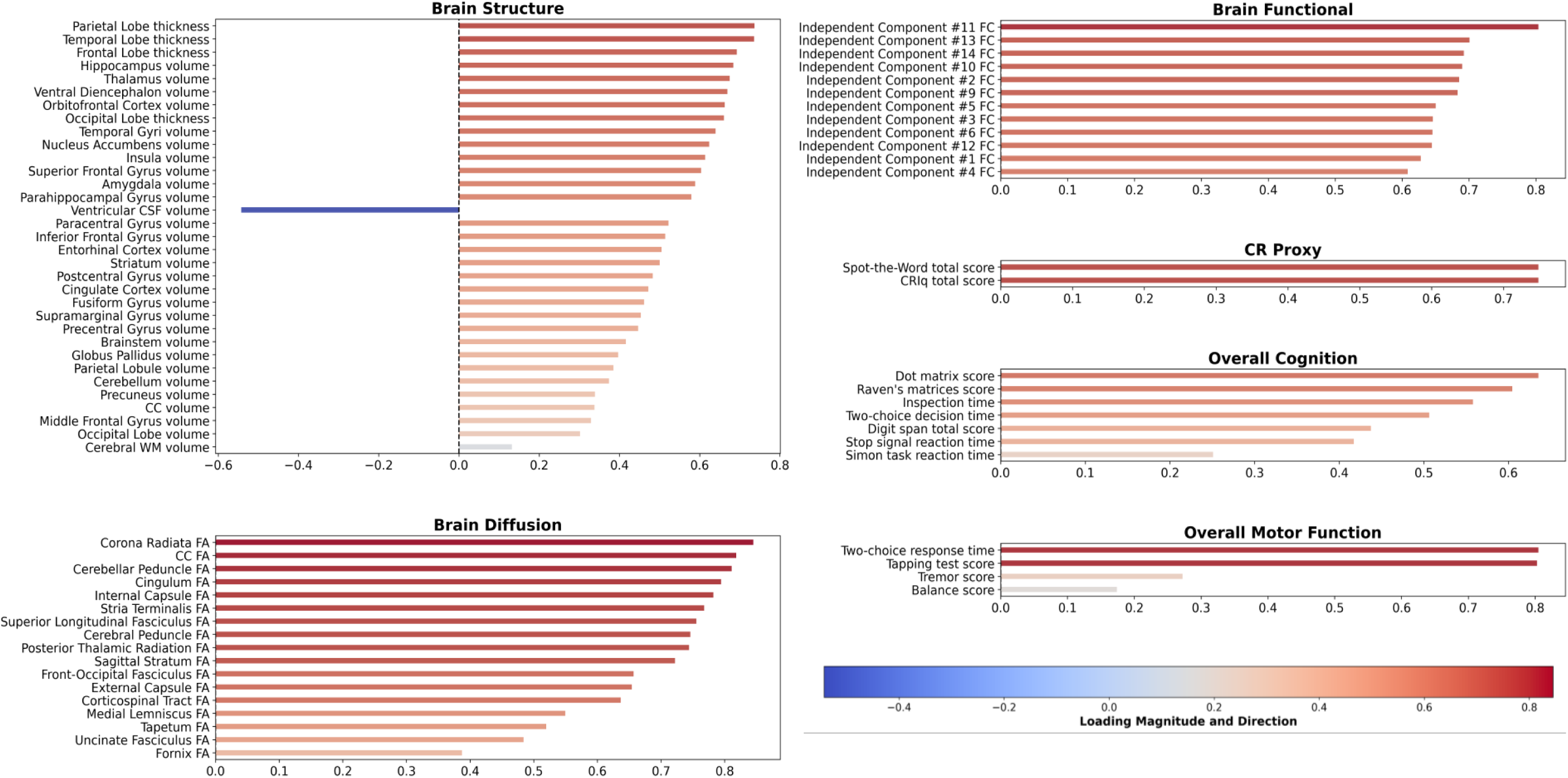
Standardised loadings for the first principal component obtained from the principal component analyses of MRI brain measures and behavioural assessments. A separate PCA was run for each imaging modality, as well as separately for cognitive reserve proxies, cognition and motor function. CC = corpus callosum. CR = cognitive reserve. CSF = cerebrospinal fluid. FA = fractional anisotropy. FC = functional connectivity. WM = white matter.

#### 2.4.1 Structural MRI analysis

T1-weighted structural images were brain extracted using FreeSurfer’s SynthStrip tool (Hoopes et al., 2022). A deep learning neuroimaging pipeline, FastSurfer (Henschel et al., 2020), was used to segment 95 brain regions in subject T1-weighted images. Masks of cerebral WM, the cerebellum, GM gyri, subcortical GM structures, the corpus callosum, brainstem, ventricular cerebrospinal fluid (CSF), and WM hypointensities were generated from FastSurfer’s parcellation. Ventricular CSF volume was chosen over peripheral CSF volume, or a combination of both, for its superior reliability in delineating between brain tissue and CSF. WM hypointensities were not included as the preferred and most reliable modality for imaging WM lesions is a T2-weighted fluid attenuated inversion recovery sequence (Duering et al., 2023). Volume of each structure was calculated using these FastSurfer segmentations, and for regions represented bilaterally, left and right hemisphere masks were combined. When anatomically sensible, brain structures were merged into one volumetric measure to reduce the number of structural brain variables used (see Supplementary Material Table 2). Mean cortical thickness for gyri in each major brain lobe (i.e., frontal, parietal, temporal, and occipital) was calculated with FastSurfer by averaging thickness metrics across structures located within lobes. All volumetric indices generated were normalised by dividing the volumes by the estimated total intracranial volume, as calculated using FastSurfer.

#### 2.4.2 Diffusion-weighted MRI analysis

Diffusion-weighted images were first corrected for susceptibility-induced distortions using FSL’s topup tool (Andersson et al., 2003), which processes pairs of images acquired with opposing phase encoding directions, but without diffusion-weighting (b=0), and then applies the derived correction to all images, including those with diffusion-weighting (b>0). All images were brain extracted before being corrected for eddy current distortions and movement, aligning them to the reference *b*=0 image, using FSL’s eddy tool (Andersson & Sotiropoulos, 2016). Following this, FSL’s DTIFIT was used to generate a fractional anisotropy (FA) image for each subject. These FA images were aligned into a common space using the nonlinear registration tool FNIRT, and a mean FA skeleton that represents the centres of all tracts common to the sample was created using TBSS (Tract-Based Spatial Statistics; Smith et al. 2006). Masks of 48 major WM tracts were created in standard space using the John Hopkins University International Consortium for Brain Mapping atlas (https://identifiers.org/neurovault.image:1401; Hua et al. 2008). To limit the number of FA measures included in analyses, left and right WM tract masks, as well as subsections of tracts, were combined (see Supplementary Material Table 3). Possible contamination from nearby GM was minimised by calculating mean FA within the WM skeleton mask generated by TBSS. Some subjects underwent a slightly different diffusion-weighted imaging (DWI) sequence (due to scanner upgrades during data collection) that generated significantly higher mean FA values. Variation across sequences was corrected by calculating the difference between the median of tract FA means for each DWI sequence to generate a tract offset value. Tract-specific offset values provided the most robust correction, effectively aligning subjects with sequence-driven elevated FA values to the rest of the sample (see Supplementary Material Fig 2).

#### 2.4.3 Resting-state functional MRI analysis

rsfMRI data was pre-processed and decomposed into different spatial and temporal components using FSL’s MELODIC (Beckmann & Smith, 2004) to perform single-subject independent component analysis (ICA). Prior to this step, the corresponding fieldmap magnitude image was brain extracted and used, alongside the fieldmap phase image, to generate a fieldmap image in radians per second for correction of distortions due to magnetic field inhomogeneities in the functional image. Functional image pre-processing steps performed within MELODIC included: motion correct via MCFLIRT (Jenkinson et al., 2002), interleaved slice timing correction, spatial smoothing with 3mm full width at half maximum, and high pass temporal filtering at 100 seconds to remove very slow drifts. Lowpass filtering was not performed since, unlike at extremely low frequencies, there can be useful neural signal at higher frequencies (Poldrack et al., 2011). For a subset of 20% of subjects, independent components (IC) output by MELODIC were manually classified as ‘Signal’ or ‘Noise’ and used to create a training file for automatic classification of ICs using FSL’s pyFIX (Salimi-Khorshidi et al., 2014) at a threshold of 20. Lower pyFIX threshold values allow more ICs to be classified as signal (even if they are noise), while higher values, like the one chosen in the current study, are stricter. Subsequently, functional images were denoised via this classification and transformed to a standard space using FSL’s applywarp. Group-level spatial maps were created by temporally concatenating filtered functional data for all subjects through group ICA in MELODIC. The short scan duration restricted group ICA dimensionality to 20. Visual inspection of all the 20 spatial maps produced revealed that there were 12 resting-state networks (RSN; see Supplementary Material Fig 3) and 8 noise components. Dual regression was performed on the 12 group-level ICs to estimate subject-specific spatial maps for each IC by regressing the group-average spatial map into each subject’s 4D space-time dataset (Nickerson et al., 2017). In addition, group-level spatial maps for RSNs were thresholded using an alternative hypothesis test based on fitting a Gaussian/gamma mixture model to the distribution of voxel intensities within a spatial map (Beckmann et al., 2005) and controlling the local false-discovery rate at *p* < 0.5 (equal balance between false positives and false negatives). A binary mask for each group-level IC was created and used to thresholded subject-specific IC spatial maps. The mean intensity of positive intensity voxels in an IC that survived thresholding was used as a measure of FC within that IC. Scanner upgrades also affected FC. To account for this, the offset between each scanner type was adjusted for by independently correcting each IC using the median intensity difference (see Supplementary Material Fig 4).

### 2.5 Lifetime experiences measurement

Lifetime experiences across three different domains (i.e., education, occupation, and leisure activities) were assessed by administering the Cognitive Reserve Index questionnaire (CRIq), a widely used, comprehensive socio-behavioural proxy measure of CR (Nucci et al., 2012). Since verbal intelligence measures also capture the contributions of a broad number of lifetime experiences (Boyle et al., 2021; Nogueira et al., 2022), a vocabulary Spot-the-Word test was also used (Baddeley et al., 1993).

### 2.6 Principal component analyses

Principal component analysis (PCA) was used to generate composite scores for all primary variables of interest by decomposing the data into orthogonal components. A separate PCA was run for each imaging modality, as well as separately for lifetime experiences, cognition and motor function. Before PCA, missing data was imputed using the median value and all data were scaled using the StandardScaler from the scikit-learn library (Pedregosa et al., 2011). Prior to analysis, the effects of age and sex were regressed out of each variable, generating residual values for MRI brain measures and behavioural assessments. These residuals formed the input to the PCA. It was expected that age would correlate strongly with each behavioural and MRI composite measure. By using a residual approach, the independent contributions of CR proxies and MRI brain measures could be explored more clearly without variance related to demographics confounding associations of interest.

Dimensionality of neuroanatomical features was reduced separately for each imaging modality (i.e., T1-weighted structural, DWI and rsfMRI) using PCA, producing a single composite per modality. For residual data from behavioural variables, a separate PCA was performed to generate a composite score for each variable type. More specifically, three summary measures were produced for lifetime experiences, cognition and motor function by using the principal component that explained the most variance (see Fig 1). To improve reliability, these PCAs were performed using the larger study dataset (*N*=1150) that included a combination of participants who did, and did not, complete an MRI. From these PCAs, scores for participants who did complete the MRI session were extracted and used as behavioural composite scores.

### 2.7 Statistical analyses

After performing PCA, statistical analyses were conducted with R v4.3.2 (R Core Team, 2022). Residual data were checked for normality by examining histograms and QQ plots. Correlations between variables were calculated for raw data and residual data. To validate if our lifetime experience composite captured the protective effect CR is suggested to confer, we evaluated the interaction between CR proxy score and MRI brain measures in regression models for overall cognition and motor function. Interaction effects for each model were converted into a semi-partial correlation coefficient (Aloe & Becker, 2012) and pooled as one overall effect using a multi-level meta-analytic technique (Viechtbauer, 2010). Interaction plots (Long 2019) and a forest plot were generated to visualise the moderating effect of lifetime experiences and validate it as a suitable CR proxy. We focused our validation analysis on structural characteristics (i.e., T1-weighted structural and DWI), as these measures are well-established indicators of brain integrity that are closely associated with reserve capacity (Stern, 2009; Stern et al., 2023). MRI brain measure composite scores were included in multiple linear regression analyses to investigate associations between brain characteristics and lifetime experiences. These MRI brain measures were also included in combination with lifetime experiences as predictors, in separate models, of overall cognition and motor function. The relaimpo package was used to calculate the relative importance of predictors in each model (Groemping, 2005). Analysis of variance (ANOVA) was used to assess whether removing either all brain measure composites simultaneously, or lifetime experiences, significantly affected explained variance in regression models of overall cognition and motor function. Similar ANOVAs were also performed for individual outcome tests.

## 3 Results

### 3.1 Principal component analyses

The first principal component from the structural MRI brain measure PCA explained 29% of the variance. Of the 33 measures, each loaded positively, and at relatively the same magnitude, onto this component, excluding ventricular CSF, which, as expected, loaded in the opposite direction as volumetric and thickness measures of brain tissue (see Fig 1). Thus, greater structural composite scores indicated greater brain volume and cortical thickness. For MRI diffusion brain measures, the first principal component explained 48% of the variance. Mean FA in each of the 17 WM tracts had positive loadings of similar magnitude onto this composite, with the fornix contributing the least (see Fig 1). A higher brain diffusion composite score corresponded with greater FA and therefore increased microstructure integrity across major WM tracts. Lastly, the rsfMRI principal component accounted for 45% of the variance in the 12 signal ICs generated from groupICA. The contributions of each IC were comparable, and loadings were all positive (see Fig 1). Greater brain functional composite scores reflected increased cortical activity associated with the RSNs across the 12 ICs.

For the composite scores produced from behavioural PCAs, greater scores were associated with more lifetime experiences (CR proxy) and better cognitive and motor function performance (see Fig 1). The CR proxy composite score produced from lifetime experience measures explained 56% of the variance and each variable loaded equally. The overall cognition composite score captured 25% of the variance and loadings of each cognitive test were relatively even. Hand dexterity assessments (tapping test and response time) were most strongly associated with the overall motor function score, which explained 35% of the variance in individual motor scores.

### 3.2 Demographics and summary statistics

Descriptive statistics for demographics and behavioural measures are provided in Table 1, and in Supplementary Material Table 4 for each brain measure included in the MRI PCAs. These summary statistics are for raw values, not residual data. Total CRIq and Spot-the-word scores positively correlated with age (CRIq = .78, *p* < .001; Spot-the-word = .52, *p* < .001), likely reflecting greater accumulation of experiences across the lifespan. Age was significantly positively correlated with a higher CR proxy score, but negatively associated with all MRI brain measure composite scores, as well as with overall cognition and motor function (see Supplementary Material Table 5). Each MRI brain measure composite had significant, positive relationships with overall cognition and motor function (see Supplementary Material Table 5); however, these did not persist after regressing out age and sex (see Supplementary Material Table 6). CR proxy score was significantly correlated with higher cognition and motor scores, even after controlling for age and sex (see Supplementary Material Table 6).

**Table 1.**
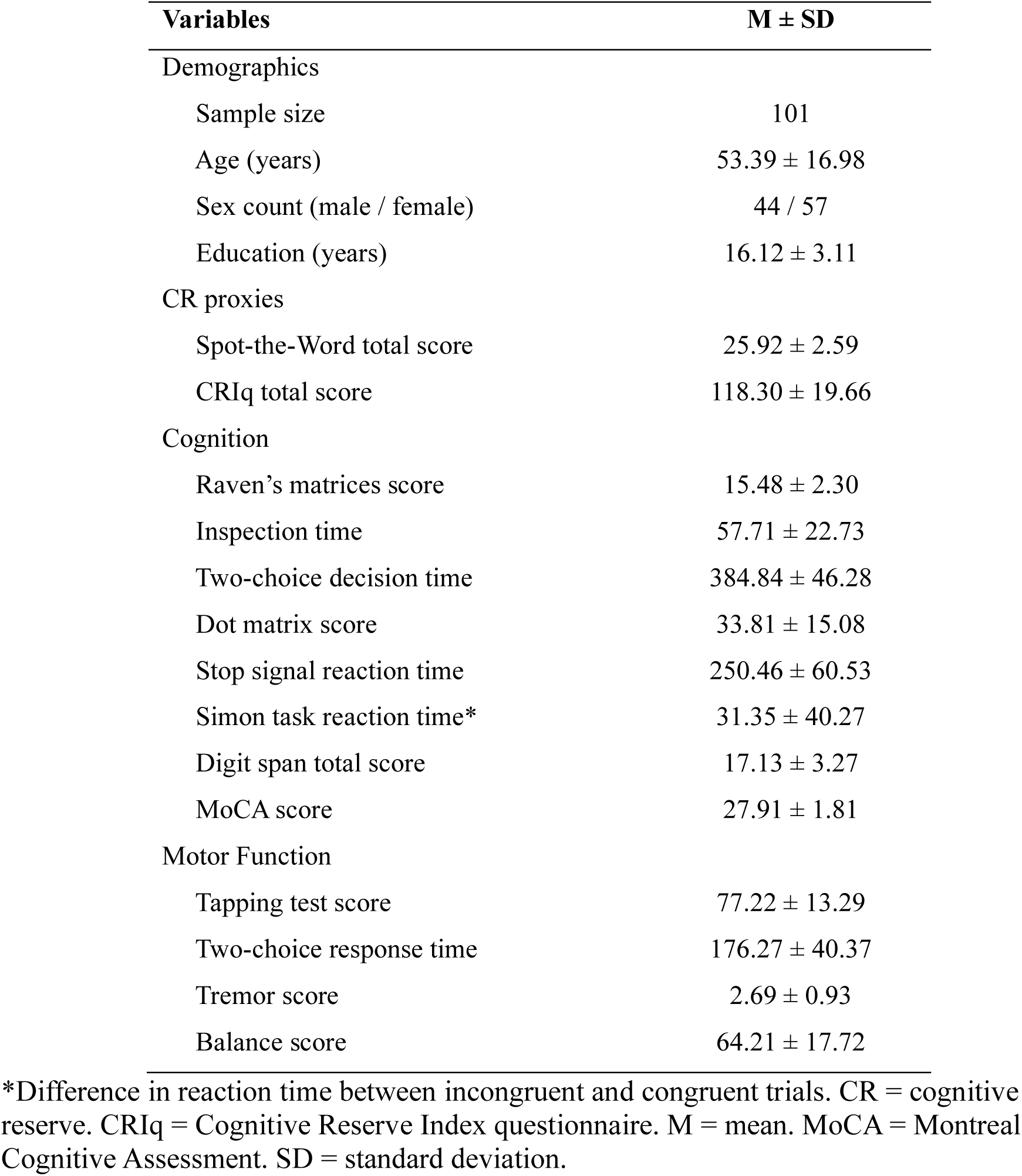
Demographic and behavioural characteristics prior to calculation of residuals and principal component analyses.

| **Variables** | **M ± SD** |
| --- | --- |
| Demographics |  |
| Sample size | 101 |
| Age (years) | 53.39 ± 16.98 |
| Sex count (male / female) | 44 / 57 |
| Education (years) | 16.12 ± 3.11 |
| CR proxies |  |
| Spot-the-Word total score | 25.92 ± 2.59 |
| CRIq total score | 118.30 ± 19.66 |
| Cognition |  |
| Raven's matrices score | 15.48 ± 2.30 |
| Inspection time | 57.71 ± 22.73 |
| Two-choice decision time | 384.84 ± 46.28 |
| Dot matrix score | 33.81 ± 15.08 |
| Stop signal reaction time | 250.46 ± 60.53 |
| Simon task reaction time* | 31.35 ± 40.27 |
| Digit span total score | 17.13 ± 3.27 |
| MoCA score | 27.91 ± 1.81 |
| Motor Function |  |
| Tapping test score | 77.22 ± 13.29 |
| Two-choice response time | 176.27 ± 40.37 |
| Tremor score | 2.69 ± 0.93 |
| Balance score | 64.21 ± 17.72 |

\*Difference in reaction time between incongruent and congruent trials. CR = cognitive reserve. CRIq = Cognitive Reserve Index questionnaire. M = mean. MoCA = Montreal Cognitive Assessment. SD = standard deviation.

### 3.3 Cognitive reserve proxy validation

The CR proxy was a significant moderator of the association between the brain structure composite score and both overall cognition and motor function (see Fig 2A). This interaction suggests that, in our cohort of healthy adults, experiences across the lifespan that presumably contribute to CR have a protective effect against structural brain alterations associated with cognitive and motor function. The CR proxy did not significantly moderate the relationship between brain diffusion composite scores and either overall cognition or motor function. Although non-significant, it is worth noting that these interactions for brain diffusion were in the expected direction. The pooled semi-partial correlation coefficient, combining interactions from both brain structure and diffusion composites, was significant, reflecting an attenuating moderation effect of lifetime experiences that is consistent with the proposed neuroprotective role of CR (see Fig 2B). Interaction plots for the brain functional composite can be viewed in Supplementary Material Fig 5.

**Fig 2.**
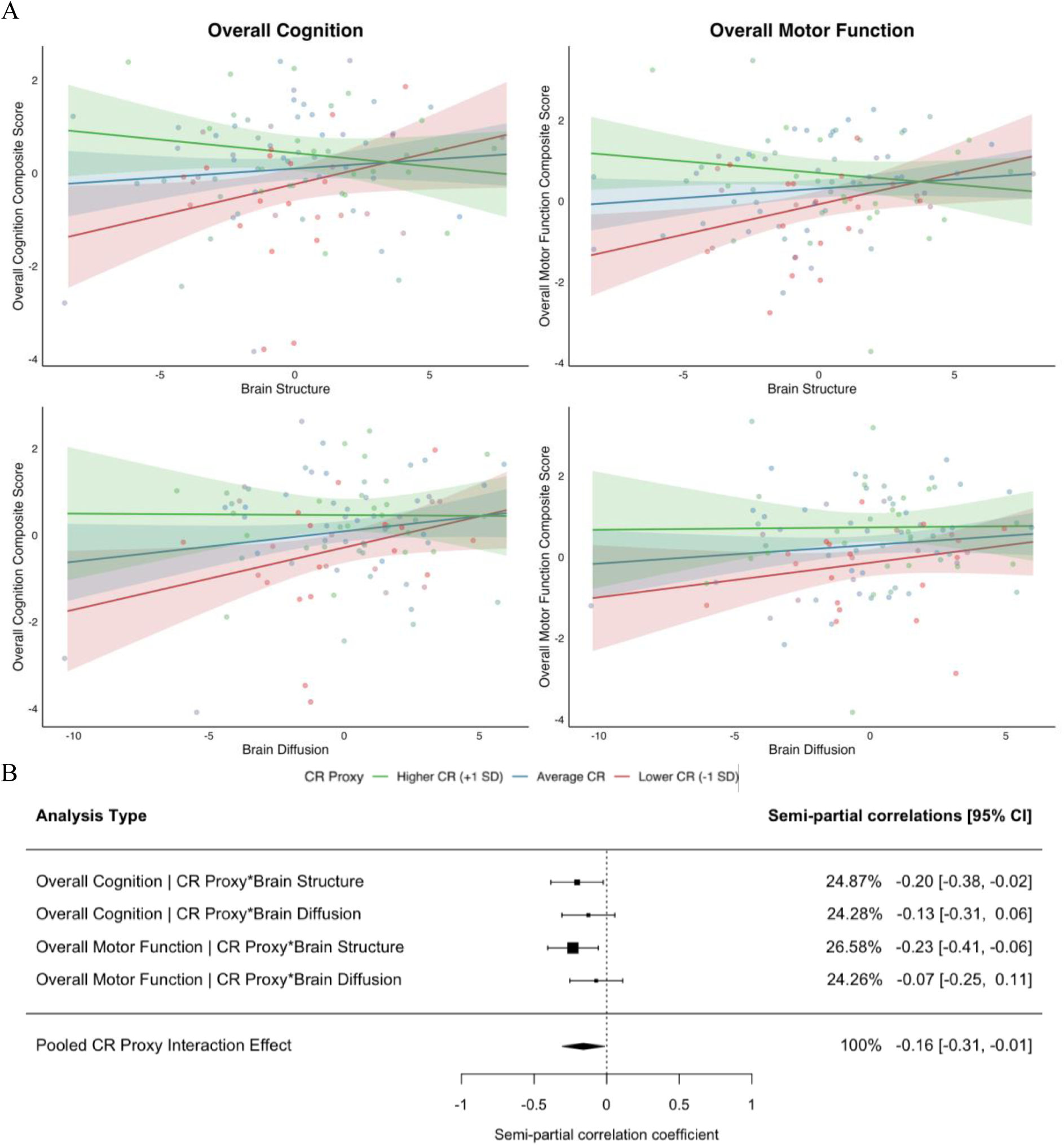
Interactions between cognitive reserve proxies and structural MRI brain measure composites for overall cognition and motor function (A). Forest plot displaying the pooled effect (semi-partial correlation coefficient) for cognitive reserve’s moderating effect (B). Higher brain measure composite scores indicated better structural health. CR = cognitive reserve.

### 3.4 Variance in cognitive reserve levels

MRI brain measures were not significantly associated with CR proxy scores, collectively explaining only ∼1% of variance after controlling for age and sex (see Table 2). This indicates that lifetime experiences were not related to brain characteristics.

**Table 2.** Results of multiple linear regressions predicting the cognitive reserve proxy and primary behavioural outcome composite scores controlling for age and sex (*p*-values ≤ .05 are shown in bold).

|  | CR proxy |  |  |  | Overall Cognition |  |  |  | Overall Motor Function |  |  |  |
| --- | --- | --- | --- | --- | --- | --- | --- | --- | --- | --- | --- | --- |
| | $\beta$ | SE | $p$ | R<sup>2</sup> | $\beta$ | SE | $p$ | R<sup>2</sup> | $\beta$ | SE | $p$ | R<sup>2</sup> |
| CR |  |  |  |  |  |  |  |  |  |  |  |  |
| CR proxy |  |  |  |  | 0.371 | 0.140 | **.010** | 0.067 | 0.418 | 0.130 | **.002** | 0.099 |
| MRI brain measures |  |  |  |  |  |  |  |  |  |  |  |  |
| Brain Structure | 0.022 | 0.031 | .466 | 0.006 | 0.002 | 0.043 | .962 | 0.003 | 0.030 | 0.039 | .454 | 0.008 |
| Brain Diffusion | 0.008 | 0.032 | .809 | 0.002 | 0.074 | 0.045 | .099 | 0.029 | 0.038 | 0.041 | .357 | 0.012 |
| Brain Functional | 0.018 | 0.039 | .640 | 0.002 | -0.007 | 0.054 | .889 | 0.000 | 0.069 | 0.050 | .169 | 0.018 |
| Total Model R<sup>2</sup> | 0.009 ($p = .831$) | | | | 0.099 ($p = \mathbf{.039}$) | | | | 0.136 ($p = \mathbf{.007}$) | | | |

$\beta$ = beta coefficient. CR = cognitive reserve. MRI = magnetic resonance imaging. $p$ = $p$ -value. R<sup>2</sup> = proportion of variance explained by predictor. SE = standard error.

### 3.5 Variance in behavioural outcomes

Behavioural outcome regression models were significant, with predictors CR proxy and MRI brain measures accounting for 10% of the variance in overall cognition and 14% of the variance in overall motor function (see Table 2). CR proxy score was associated with overall cognition beyond MRI brain measures, explaining 7% of the variance. Of the MRI measures, brain diffusion accounted for the most variance in overall cognition (see Fig 3); however, all MRI measures were all non-significant predictors. CR proxy score was also a significant predictor of overall motor function beyond MRI brain measures, explaining 10% of the variance (see Table 2). None of the three MRI brain measures were significantly associated with overall motor function and each explained minimal variance (see Fig 3).

**Fig 3.**
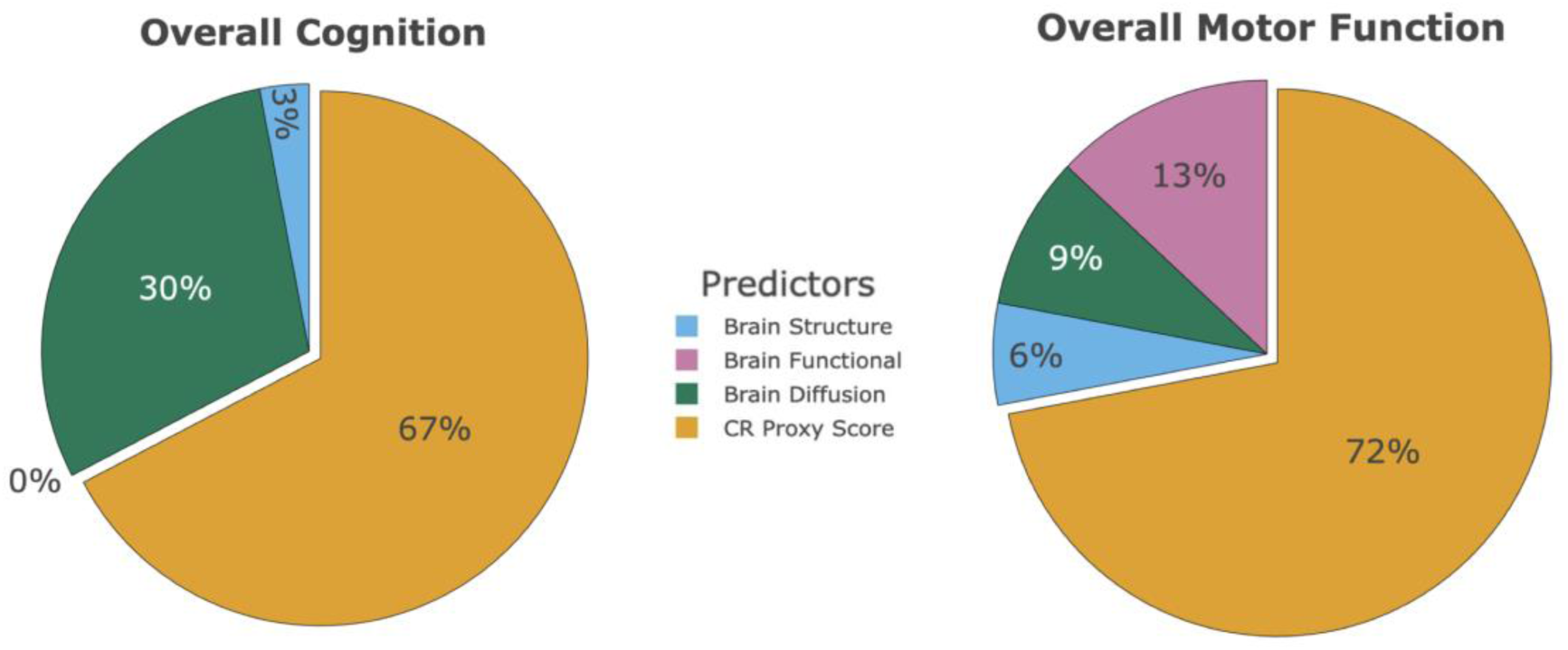
Relative importance pie charts showing the distribution of the total explained variance by significant predictors in primary behavioural outcome regression models. CR = cognitive reserve.

### 3.6 Behavioural outcome variance explained by cognitive reserve proxies compared to brain characteristics

In the ANOVAs comparing models that included either CR proxy scores or MRI brain measures vs models including both predictor types, participants with cognitive or motor outcome scores outside the expected range for a healthy adult (exceeding three standard deviations from the mean) were excluded. A single participant was excluded from each of the tremor (n=100) and balance (n=100) models, involving different individuals in each case (see Table 3). Removing the CR proxy from these models significantly reduced the total variance explained in outcomes: Raven’s matrices, inspection time, MoCA, overall cognition, tapping test, two-choice response time, and overall motor function (see Table 3). The variance accounted for by the CR proxy score was higher than the variance explained by the three MRI brain measures combined for three out of eight individual cognitive tests (Raven’s matrices score; digit span total score; MoCA score) (see Supplementary Material 7), as well as for the overall cognition composite score. Similarly, the variance explained by the CR proxy score was greater than that explained by the three brain MRI measures for all individual motor function tests (see Supplementary Table 8), as well as for overall motor function composite score. In these models, the CR proxy’s greater explained variance in outcome performance was especially noteworthy given that it, as a single predictor, outperformed all three MRI brain measures combined.

**Table 3.**
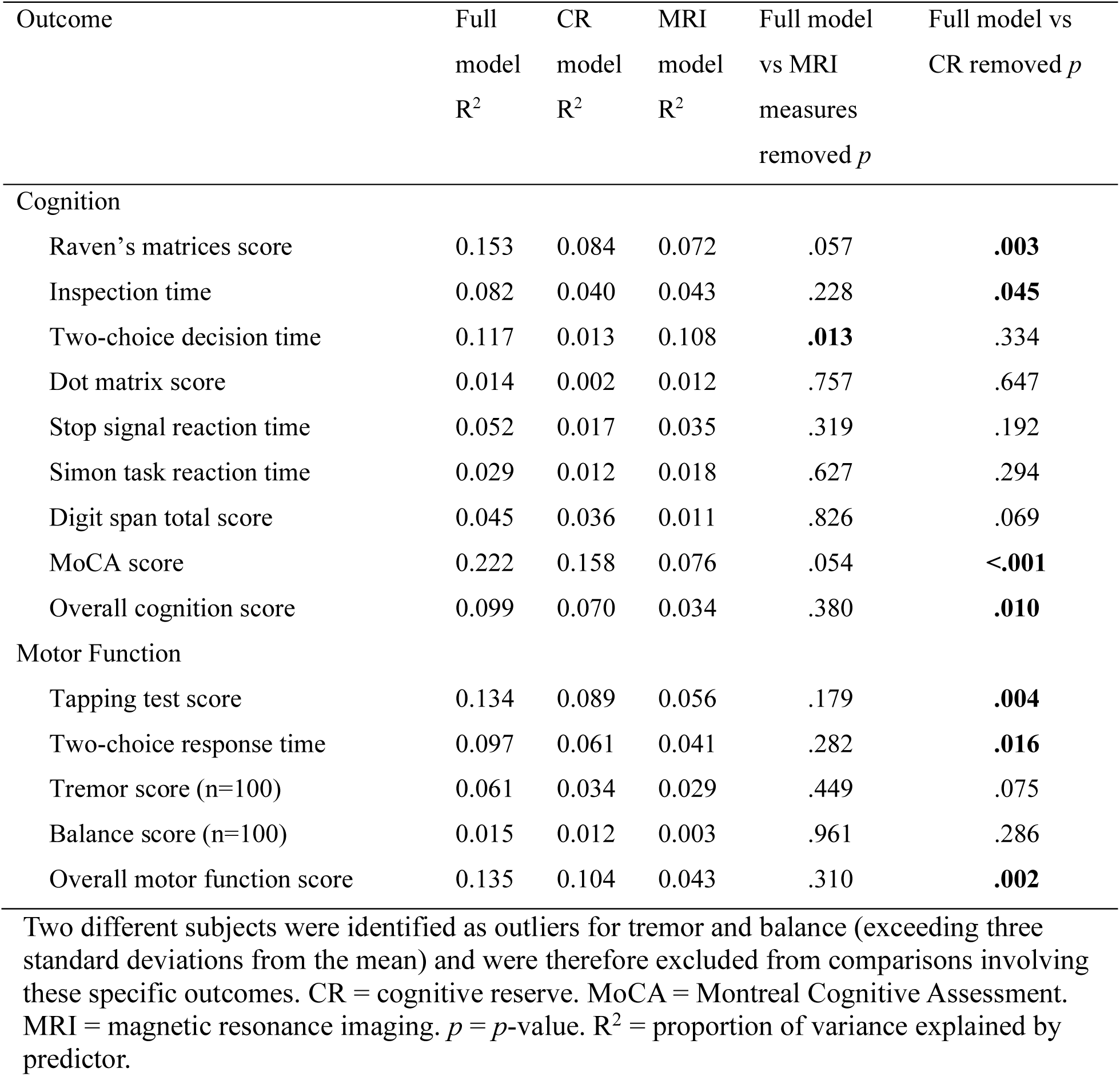
Analysis of variance results for each individual outcome score, comparing variance explained by regression models when either only the CR proxy or all MRI brain measures are dropped from the full model (uncorrected significant p-values ≤ .05 are shown in bold).

| Outcome | Full<br>model<br>$R^2$ | CR<br>model<br>$R^2$ | MRI<br>model<br>$R^2$ | Full model<br>vs MRI<br>measures<br>removed $p$ | Full model vs<br>CR removed $p$ |
| --- | --- | --- | --- | --- | --- |
| **Cognition** |  |  |  |  |  |
| Raven's matrices score | 0.153 | 0.084 | 0.072 | .057 | **.003** |
| Inspection time | 0.082 | 0.040 | 0.043 | .228 | **.045** |
| Two-choice decision time | 0.117 | 0.013 | 0.108 | **.013** | .334 |
| Dot matrix score | 0.014 | 0.002 | 0.012 | .757 | .647 |
| Stop signal reaction time | 0.052 | 0.017 | 0.035 | .319 | .192 |
| Simon task reaction time | 0.029 | 0.012 | 0.018 | .627 | .294 |
| Digit span total score | 0.045 | 0.036 | 0.011 | .826 | .069 |
| MoCA score | 0.222 | 0.158 | 0.076 | .054 | **<.001** |
| Overall cognition score | 0.099 | 0.070 | 0.034 | .380 | **.010** |
| **Motor Function** |  |  |  |  |  |
| Tapping test score | 0.134 | 0.089 | 0.056 | .179 | **.004** |
| Two-choice response time | 0.097 | 0.061 | 0.041 | .282 | **.016** |
| Tremor score (n=100) | 0.061 | 0.034 | 0.029 | .449 | .075 |
| Balance score (n=100) | 0.015 | 0.012 | 0.003 | .961 | .286 |
| Overall motor function score | 0.135 | 0.104 | 0.043 | .310 | **.002** |

Two different subjects were identified as outliers for tremor and balance (exceeding three standard deviations from the mean) and were therefore excluded from comparisons involving these specific outcomes. CR = cognitive reserve. MoCA = Montreal Cognitive Assessment. MRI = magnetic resonance imaging. $p$ = $p$ -value. $R^2$ = proportion of variance explained by predictor.

MRI brain measures explained a higher proportion of variance compared to CR proxy scores in five out of eight cognitive outcomes: inspection time; two-choice decision time; dot matrix score; SSRT; and Simon task reaction time (see Table 3). For most of these outcomes, where MRI brain measures explained more variance, CR proxy score explained little to no variance, excluding inspection time, where it was a significant predictor (see Supplementary Material Table 7). This effect was most pronounced for the two-choice decision time, which was the only outcome to show a significant reduction in explained variance when MRI brain measures were removed from the model (see Table 3).

## 4 Discussion

CR theory postulates that lifetime experiences contribute to underlying differences in cognitive processing that enable more successful task completion, regardless of differences in neuroanatomical features associated with these tasks (Stern, 2002). Thus, CR should account for variance in task performance that cannot be attributed to any brain characteristics. While past studies have outlined possible biological substrates of CR, they have not yet determined the variance in behavioural outcomes that lifetime experiences, as a CR proxy, can account for beyond brain measures. Here, to the best of our knowledge, we explore this for the first time using a multimodal MRI approach. Our measure of lifetime experiences was validated as a proxy for CR by demonstrating that it moderates the association between structural brain indices and behavioural outcomes in healthy adults. The current study showed that neither structural, diffusion, or functional MRI brain measure composites explained a significant amount of variance in CR proxies. Interestingly, CR proxies were a significant predictor of both overall cognitive and motor function, while brain measures did not predict either outcome. Further, the relative importance of CR proxies was greater for outcome assessments of specific higher-level cognitive abilities and individual motor tests evaluating voluntary movements.

### 4.1 Cognitive reserve proxy validation

Although not the study’s primary focus, we aimed to validate our lifetime experiences composite as a proxy of CR in healthy adults without pathological brain changes. Significant interactions were observed between CR proxy score and the MRI brain structure composite score for both overall cognition and motor function. This aligns with previously reported moderation effects of protective lifetime experiences in healthy adults, both in studies examining brain structure (Jin et al., 2023; Querbes et al., 2009; Stern et al., 2018; Vuoksimaa et al., 2013) and in those investigating markers of typical age-related pathology (Brickman et al., 2011; Rentz et al., 2010). Interestingly, interactions between CR proxies and the brain composite derived from diffusion measures were non-significant for both outcomes in the current study, although they trended in the expected direction. Prior work, using a large UK Biobank sample, identified significant interactions between CR proxies and WM tract FA, though these were rare across the many models performed (Lin et al., 2023). This highlights the difficulties associated with demonstrating the significant moderating effects of lifetime experiences as evidence of CR unless there is extensive statistical power available (Boyle et al., 2021). Nevertheless, the pooled interaction with all structural brain composite measures was significant, validating our lifetime experience measure as a proxy of CR.

### 4.2 Variance in cognitive function

In our sample of healthy adults, CR proxy score was the sole significant predictor of overall cognition. The positive relationship between CR proxies and overall cognition is consistent with findings from previous studies of healthy adults (Opdebeeck et al., 2016; Panico et al., 2022). Crucially, our analysis indicated that CR proxies explain individual differences in cognition beyond what can be explained by structural, diffusion or functional brain characteristics. Interestingly, none of the MRI brain measure composite scores accounted for a significant portion of variance in overall cognition, despite using a multimodal approach. This suggests that enriching lifetime experiences influence cognitive performance through mechanisms that are not captured by structural, diffusion or functional MRI in healthy adults. For instance, in individuals without neurodegeneration, lifetime experiences that impart CR are thought to underlie adaptability in cognitive processes through greater neural efficiency, capacity, or flexibility, which maximise task performance, despite brain status (Barulli & Stern, 2013; Tucker & Stern, 2011). That is, individual differences in healthy adult cognition are more likely to reflect variation in CR-related neural dynamics that lifetime experiences grant, which MRI brain measures might not account for if these changes occur at the cellular level. These findings support the notion that CR confers cognitive advantages, even in the absence of detectable neuroanatomical differences or pathological progression.

CR theory was developed according to the findings that enriching lifetime experiences enhance general cognitive ability (Stern, 2009). Consistent with this, CR proxies showed stronger associations with global cognition than with specific cognitive domains in healthy older adults (Jin et al., 2023; Lavrencic et al., 2018). Our findings align with this pattern, suggesting that CR operates broadly across cognitive networks to preserve overall function, rather than enhancing isolated domain-specific abilities. CR proxy score as a single predictor explained significantly more variance than all three MRI brain measures combined in multiple global cognitive measures (Raven’s matrices; MoCA; overall cognition composite) that test several cognitive abilities. Greater reasoning ability test performance, such as is measured by the Raven’s matrices test, has been previously associated with superior reading comprehension (Cockcroft & Israel, 2011; Tucker & Stern, 2011) and more years of education (Tucker-Drob et al., 2009), suggesting that a higher CR may enable more efficient use of neural resources to help perform optimally in day-to-day tasks. Further, removing all MRI brain measures from global cognitive test models did not significantly reduce the explained variance, but there was an unexpected significant negative association between the brain functional composite and Raven’s matrices. However, CR proxy score was a stronger predictor, explaining almost twice the variance in Raven’s matrices scores.

Although the MoCA is primarily used to detect cognitive impairment, it may also be a useful assessment for identifying global cognitive differences in healthy adults (Bruijnen et al., 2020; Gluhm et al., 2013). Removing CR proxies from the MoCA model led to a significant reduction in explained variance. Additionally, the brain diffusion composite, independent of other MRI brain measures, was a significant predictor of MoCA performance, such that higher FA was associated with better MoCA scores. These findings likely reflect the multifactorial nature of the MoCA (Nasreddine et al., 2005), where specific subtests could be influenced by either lifetime experiences or brain microstructure, or the combination of the two, whereas the overall score appears to be influenced by both. In line with this, prior healthy adult studies have linked MoCA performance to brain diffusivity (Jolly et al., 2016; Wei et al., 2019), as well as differences in education years and occupational complexity (Liu et al., 2013; Montemurro et al., 2023). To our knowledge, this study is the first to show that both CR proxies and FA independently contribute to global cognition, indicating that the MoCA may be sensitive for detecting differences in neuropsychological performance differences even among cognitively healthy adults, and not only in individuals with cognitive impairment.

In contrast, more basic cognitive abilities, such as visuomotor processing speed (Ritchie et al., 2013), which are among the first to decline with age-related pathology (Salthouse, 2009), are less likely to be preserved by lifetime experiences (Lavrencic et al., 2018). Our results partly suggest a similar pattern. The low relative importance attributed to CR proxies in our models of individual cognitive measures including two-choice decision time, SSRT, dot matrix and Simon task, provides further evidence for this claim. Furthermore, we found that the brain diffusion composite was typically the strongest MRI brain measure predictors of processing speed tasks, and often stronger than CR proxy score. The relationship we observed between WM integrity and processing speed is consistent with established findings in the broader literature (Bennett & Madden, 2014; Kerchner et al., 2012; Madden et al., 2012). However, CR proxy score was a significant predictor of inspection time, and removing it from the model significantly reduced the explained variance, suggesting a potential influence of lifetime experiences on information processing mechanisms. As CR proxy score did not predict other basic cognitive abilities, this effect warrants further investigation, and our conclusions here remain tentative.

### 4.3 Variance in motor function

A higher CR proxy score was also associated with better overall motor function, beyond MRI brain measures. Removing lifetime experiences, but not MRI brain measures, resulted in significant reductions in the variance explained by the model. These findings suggest that lifetime experiences explain variance in motor function test performance that differences in brain characteristics are less sensitive to, similar to what we observed for overall cognition. To our knowledge, the association between CR proxies and motor function has been largely under-investigated. In our study, overall cognition and motor function correlated highly. This is not surprising, since some motor tasks required cognitive planning and execution of voluntary movements (Kobayashi-Cuya et al., 2018; Leisman et al., 2016). Normal ageing studies suggest an association between cognitive and motor function, given that both of these abilities are affected by neural changes, such as reduced brain volume or connectivity (Clouston et al., 2013; Kong et al., 2020; Sánchez-Izquierdo & Fernández-Ballesteros, 2021). Thus, if CR proxies can predict differences in cognitive abilities, then they should also account for variation in motor outcomes. The reliance on higher-level cognitive abilities for successful movements may explain why the effect of CR proxies indirectly extend to motor function.

Interestingly, CR proxies explained more variance in motor tasks involving voluntary movements, such as the tapping test score and two-choice response time, compared to those assessing involuntary movements (i.e. balance and tremor). Similarly, removing CR significantly impacted model performance for both tasks. This may reflect the stronger cognitive demands of voluntary actions required in the tapping test and two-choice response time task. Given the established association between cognitive and motor function (Kobayashi-Cuya et al., 2018; Leisman et al., 2016), and evidence that hand dexterity predicts executive function (Kobayashi-Cuya et al., 2024), it is perhaps unsurprising that lifetime experiences influencing cognition also affect voluntary motor performance. However, more evidence acquired from studies examining the predictive value of lifetime experiences for tasks involving voluntary movements is required to establish stronger conclusions about the role of CR in optimising and preserving motor function.

### 4.4 No association between brain characteristics and cognitive reserve proxies

Notably, no MRI brain measure composite was associated with CR proxy score, after controlling for age and sex, in this study. Strong relationships between CR proxies and brain structure have been extensively reported previously. Accumulation of more enriching lifetime experiences have been associated with greater total brain volume (Devita et al., 2024; Valenzuela et al., 2008; Yang et al., 2024), increased cortical thickness (Liu et al., 2012) and lower CSF volume (Coffey et al., 1999). Furthermore, despite prior work demonstrating relationships between lifetime experiences and RSN FC (Bozzali et al., 2015; Conti et al., 2021; Franzmeier et al., 2017; Montemurro et al., 2023; Varela-López et al., 2022; Weiler et al., 2018; Ye et al., 2022), our study also found no associations between our CR proxy score and the composite for RSN-related brain activity. While these previous studies generally focused on mid- to late-life healthy adults, we investigated CR across a wider age range to assess whether it confers advantages across the broader adult lifespan. The inclusion of younger adults may have reduced age-related variability in MRI brain measures, potentially contributing to the absence of detectable relationships with the CR proxy score. Importantly, however, the current study demonstrated that the effect of lifetime experiences on both cognitive and motor function occurs beyond brain characteristics and other covariates. Together with these findings, the non-significant relationship between our CR proxy and each MRI brain measure further suggests that the influence of lifetime experiences on healthy adult task performance may well operate independently of structural and functional brain properties that can be captured by MRI.

### 4.5 Limitations and future directions

There are some limitations that should be considered when interpreting these findings. Our sample size was relatively modest; however, it was comparable to previous analyses of CR proxies and MRI brain measures in healthy adults (Conti et al., 2021), and sufficient to detect a robust effect of lifetime experiences that was independent of brain characteristics, age and sex. Future larger studies with adequate power to test age group differences would be useful to provide additional insight into the independent effects of CR across the lifespan.

Moreover, in our study, CR was evaluated using a composite score, as it generates a better estimation of the construct (Grotz et al., 2017). It is worth noting, however, that analysing the effects of individual proxies may provide additional information regarding CR’s influence. Indeed, early-life CR proxies (i.e., education years) are crucial for maintaining cognitive performance, but one study demonstrated that late-life proxies (i.e., social engagement) may be more important for preventing cognitive decline in late life (Yang et al., 2022).

Furthermore, composite measures were created to preserve statistical power and reduce the number of comparisons, while still accounting for contributions from several neuroanatomical features and outcome domains. Previous studies have reported associations between CR proxies and individual structural and functional brain measures (Conti et al., 2021; Yang et al., 2024), as well as links between specific brain regions and particular outcome variables (Fjell & Walhovd, 2010; Raz et al., 2005). Although beyond the scope of the current study, future research may benefit from exploring the relationships among different CR proxies, independent brain measures, and individual outcome tests associated with these specific neuroanatomical characteristics.

## 5 Conclusion

Our study was the first to demonstrate that lifetime experiences, as a CR proxy, independently predict both cognitive and motor function in healthy adults, beyond structural and functional brain characteristics. This finding suggests that CR extends beyond mitigating detrimental age-related brain changes, enhancing brain processes that support superior cognition and motor function in the absence of pathology. Further, we observed that lifetime experiences attenuated the association between structural brain characteristics and both overall cognition and motor function. Consequently, participating in stimulating lifetime experiences that contribute to CR not only compensates for neurobiological decline but also increases task performance across the lifespan, thereby helping maximise quality of life throughout adulthood. These results highlight the value of integrating both CR and neuroimaging measures to better predict task performance, while emphasising the importance of promoting lifelong engagement in stimulating experiences to enhance and optimise behavioural outcomes.

## List of abbreviations

ANOVA: Analysis of Variance
CR: Cognitive Reserve
CRIq: Cognitive Reserve Index questionnaire
CSF: Cerebrospinal Fluid
DTIFIT: Diffusion Tensor Imaging Fit
DWI: Diffusion-weighted Imaging
FA: Fractional Anisotropy
FC: Functional Connectivity
FNIRT: FMRIB’s Non-linear Image Registration Tool
FSL: FMRIB Software Library
GM: Grey Matter
IC: Independent Component
ICA: Independent Component Analysis
MCFLIRT: Motion Correction using FMRIB’s Linear Image Registration Tool
MELODIC: Multivariate Exploratory Linear Optimization for Decomposition of Independent Components
MoCA: Montreal Cognitive Assessment
MRI: Magnetic Resonance Imaging
PCA: Principal Component Analysis
pyFIX: Python implementation of functional MRI Independent Component Extraction X-classifier
rsfMRI: Resting-state Functional Magnetic Resonance Imaging
RSN: Resting-state Network
SSRT: Stop Signal Reaction Time
TBSS: Tract-based Spatial Statistics
WM: White Matter

## Declaration of Competing Interest

The authors declare no competing interests and have nothing to disclose.

## Funding

This work was supported by grants to LCP, MJ, IB, AD, and AW from the Medical Research Future Fund (2020/MRF1202188; Forecasting Impairment and Neurodegenerative Disease risk following Traumatic Brain Injury (FIND-TBI): A computational neurology-driven method to predict long-term prognosis) and the Australian Research Council Discovery Project to IB and LCP (DP190103600; Investigating differences in decision-making ability in older adults: Computational modelling and neurogenetics of the basal ganglia). IS was supported by the University of Adelaide Research Scholarship. BC was supported by an Australian Government Research Training Program Scholarship.

## Data Availability Statement

Due to the current study using a subset of data from a larger project, the dataset used in this study is not publicly available. However, upon completion of the larger project, all data collected will be made widely accessible. In the meantime, the data used in this study may be made available upon request.

## Authorship contribution statement (CRediT)

**Isaac Saywell:** Writing – review & editing, Writing – original draft, Methodology, Visualisation, Investigation, Formal analysis, Data curation, Conceptualisation. **Sabrina Sghirripa:** Writing – review & editing, Methodology, Investigation, Formal analysis, Conceptualisation. **Angela Walls:** Writing – review & editing, Funding acquisition. **Andrew Dwyer:** Writing – review & editing, Funding acquisition. **Brittany Child:** Writing – review & editing, Data curation, Visualisation, Software. **John Salamon:** Writing – review & editing, Visualisation, Software. **Lyndsey Collins-Praino:** Writing – review & editing, Supervision, Funding acquisition, Project administration, Conceptualisation. **Mark Jenkinson:** Writing – review & editing, Supervision, Funding acquisition, Methodology, Investigation, Project administration, Software, Formal analysis, Conceptualisation. **Irina Baetu:** Writing – review & editing, Supervision, Funding acquisition, Methodology, Investigation, Project administration, Formal analysis, Conceptualisation.

## Supplementary material

Supplementary material associated with this article can be found in the online version of the article.

## Acknowledgements

We acknowledge all study participants and research assistants who contributed to data collection. The authors acknowledge the Software from the University of Minnesota Center for Magnetic Resonance Research. The authors would also like to acknowledge the facilities and scientific and technical assistance of the National Imaging Facility, a National Collaborative Research Infrastructure Strategy (NCRIS) capability.

## Notes

### Competing Interest Statement

The authors have declared no competing interest.

### Author Declarations

Ethics approval was obtained from the University of Adelaide Human Research Ethics Committee (approval numbers H-2021-120 and H-2020-017)

### Summary of Updates

The manuscript has been revised to update the methodology and results. The original analysis performed dimensionality reduction on all MRI brian measures in the same model. However, the updated manuscript analyses each MRI modality independently, improving the interpretability of the data. Further, to keep the manuscript focused on functional outcomes mood was removed, meaning that only cognition and motor function were evaluated. To ensure that our lifetime experience composite was validated as a proxy of CR, and therefore could be interpreted as so, we performed interaction analyses. These analyses involved assessing whether CR moderated the relationship between composite structural brain measures and both cognition and motor function. These methodological changes helped improve the ability to interpret what MRI brain measure composites represented, and made the final results clearer. Thus, the results and interpretation of these findings in the discussion section have changed to accomodate these methodological adjustments. The original aims and hypotheses of the manuscript remain the same. The format of some figures and tables were adjusted to improve clarity and interpretability of findings. Supplementary material have also been updated to represent these methodological changes.

## References

Aloe, A. M., & Becker, B. J. (2012). An effect size for regression predictors in meta-analysis. Journal of Educational and Behavioral Statistics, 37, 278–297. 10.3102/1076998610396901

Anders M. Fjell, & Kristine B. Walhovd. (2010). Structural Brain Changes in Aging: Courses, Causes and Cognitive Consequences. Reviews in the Neurosciences, 21(3), 187–222. 10.1515/REVNEURO.2010.21.3.187

Andersson, J. L. R., Skare, S., & Ashburner, J. (2003). How to correct susceptibility distortions in spin-echo echo-planar images: Application to diffusion tensor imaging. NeuroImage, 20(2), 870–888. 10.1016/S1053-8119(03)00336-7

Andersson, J. L. R., & Sotiropoulos, S. N. (2016). An integrated approach to correction for off-resonance effects and subject movement in diffusion MR imaging. NeuroImage, 125, 1063–1078. 10.1016/j.neuroimage.2015.10.019

Arenaza-Urquijo, E. M., Bosch, B., Sala-Llonch, R., Solé-Padullés, C., Junqué, C., Fernández-Espejo, D., Bargalló, N., Rami, L., Molinuevo, J. L., & Bartrés-Faz, D. (2011). Specific Anatomic Associations Between White Matter Integrity and Cognitive Reserve in Normal and Cognitively Impaired Elders. The American Journal of Geriatric Psychiatry, 19(1), 33–42. 10.1097/JGP.0b013e3181e448e1

Baddeley, A., Emslie, H., & Nimmo-Smith, I. (1993). The Spot-the-Word test: A robust estimate of verbal intelligence based on lexical decision. British Journal of Clinical Psychology, 32(1), 55–65. 10.1111/j.2044-8260.1993.tb01027.x

Barulli, D., & Stern, Y. (2013). Efficiency, capacity, compensation, maintenance, plasticity: Emerging concepts in cognitive reserve. Trends in Cognitive Sciences, 17(10), 10.1016/j.tics.2013.08.012. https://doi.org/10.1016/j.tics.2013.08.012

Beckmann, C. F., DeLuca, M., Devlin, J. T., & Smith, S. M. (2005). Investigations into resting-state connectivity using independent component analysis. Philosophical Transactions of the Royal Society of London. Series B, Biological Sciences, 360(1457), 1001–1013. 10.1098/rstb.2005.1634

Beckmann, C. F., & Smith, S. M. (2004). Probabilistic independent component analysis for functional magnetic resonance imaging. IEEE Transactions on Medical Imaging, 23(2), 137–152. IEEE Transactions on Medical Imaging. 10.1109/TMI.2003.822821

Bennett, I. J., & Madden, D. J. (2014). Disconnected aging: Cerebral white matter integrity and age-related differences in cognition. Neuroscience, 276, 187–205. 10.1016/j.neuroscience.2013.11.026

Boyle, R., Knight, S. P., De Looze, C., Carey, D., Scarlett, S., Stern, Y., Robertson, I. H., Kenny, R. A., & Whelan, R. (2021). Verbal intelligence is a more robust cross-sectional measure of cognitive reserve than level of education in healthy older adults. Alzheimer’s Research & Therapy, 13(1), 128. 10.1186/s13195-021-00870-z

Bozzali, M., Dowling, C., Serra, L., Spanò, B., Torso, M., Marra, C., Castelli, D., Dowell, N. G., Koch, G., Caltagirone, C., & Cercignani, M. (2015). The impact of cognitive reserve on brain functional connectivity in Alzheimer’s disease. Journal of Alzheimer’s Disease: JAD, 44(1), 243–250. 10.3233/JAD-141824

Brichko, R., Soldan, A., Zhu, Y., Wang, M.-C., Faria, A., Albert, M., Pettigrew, C., & The BIOCARD Research Team. (2022). Age-Dependent Association Between Cognitive Reserve Proxy and Longitudinal White Matter Microstructure in Older Adults. Frontiers in Psychology, 13. 10.3389/fpsyg.2022.859826

Brickman, A. M., Siedlecki, K. L., Muraskin, J., Manly, J. J., Luchsinger, J. A., Yeung, L.-K., Brown, T. R., DeCarli, C., & Stern, Y. (2011). White matter hyperintensities and cognition: Testing the reserve hypothesis. Neurobiology of Aging, 32(9), 1588–1598. 10.1016/j.neurobiolaging.2009.10.013

Bruijnen, C. J. W. H., Dijkstra, B. A. G., Walvoort, S. J. W., Budy, M. J. J., Beurmanjer, H., De Jong, C. A. J., & Kessels, R. P. C. (2020). Psychometric properties of the Montreal Cognitive Assessment (MoCA) in healthy participants aged 18–70. International Journal of Psychiatry in Clinical Practice, 24(3), 293–300. 10.1080/13651501.2020.1746348

Cai, Y., Tian, Q., Gross, A. L., Wang, H., E, J.-Y., Agrawal, Y., Simonsick, E. M., Ferrucci, L., & Schrack, J. A. (2022). Motor and Physical Function Impairments as Contributors to Slow Gait Speed and Mobility Difficulty in Middle-Aged and Older Adults. The Journals of Gerontology. Series A, Biological Sciences and Medical Sciences, 77(8), 1620–1628. 10.1093/gerona/glac001

Clark, R. A., Bryant, A. L., Pua, Y., McCrory, P., Bennell, K., & Hunt, M. (2010). Validity and reliability of the Nintendo Wii Balance Board for assessment of standing balance. Gait & Posture, 31(3), 307–310. 10.1016/j.gaitpost.2009.11.012

Clouston, S. A. P., Brewster, P., Kuh, D., Richards, M., Cooper, R., Hardy, R., Rubin, M. S., & Hofer, S. M. (2013). The Dynamic Relationship Between Physical Function and Cognition in Longitudinal Aging Cohorts. Epidemiologic Reviews, 35(1), 33–50. 10.1093/epirev/mxs004

Cockcroft, K., & Israel, N. (2011). The Raven’s Advanced Progressive Matrices: A Comparison of Relationships with Verbal Ability Tests. South African Journal of Psychology, 41(3), 363–372. 10.1177/008124631104100310

Coffey, C. E., Saxton, J. A., Ratcliff, G., Bryan, R. N., & Lucke, J. F. (1999). Relation of education to brain size in normal aging. Neurology, 53(1), 189–189. 10.1212/WNL.53.1.189

Conti, L., Riccitelli, G. C., Preziosa, P., Vizzino, C., Marchesi, O., Rocca, M. A., & Filippi, M. (2021). Effect of cognitive reserve on structural and functional MRI measures in healthy subjects: A multiparametric assessment. Journal of Neurology, 268(5), 1780– 1791. 10.1007/s00415-020-10331-6

Damoiseaux, J. S., Beckmann, C. F., Arigita, E. J. S., Barkhof, F., Scheltens, P., Stam, C. J., Smith, S. M., & Rombouts, S. a. R. B. (2008). Reduced resting-state brain activity in the “default network” in normal aging. Cerebral Cortex (New York, N.Y.: 1991), 18(8), 1856–1864. 10.1093/cercor/bhm207

Daneault, J.-F., Carignan, B., Codère, C. É., Sadikot, A. F., & Duval, C. (2013). Using a Smart Phone as a Standalone Platform for Detection and Monitoring of Pathological Tremors. Frontiers in Human Neuroscience, 6, 357. 10.3389/fnhum.2012.00357

Devita, M., Debiasi, G., Anglani, M., Ceolin, C., Mazzonetto, I., Begliomini, C., Cauzzo, S., Raffaelli, C., Lazzarin, A., Ravelli, A., Bordignon, A., De Rui, M., Sergi, G., Bertoldo, A., Mapelli, D., & Coin, A. (2024). The Role of Cognitive Reserve in Protecting Cerebellar Volumes of Older Adults with mild Cognitive Impairment. Cerebellum (London, England). 10.1007/s12311-024-01695-w

Duering, M., Biessels, G. J., Brodtmann, A., Chen, C., Cordonnier, C., Leeuw, F.-E. de, Debette, S., Frayne, R., Jouvent, E., Rost, N. S., Telgte, A. ter, Salman, R. A.-S., Backes, W. H., Bae, H.-J., Brown, R., Chabriat, H., Luca, A. D., deCarli, C., Dewenter, A.,… Wardlaw, J. M. (2023). Neuroimaging standards for research into small vessel disease—Advances since 2013. The Lancet Neurology, 22(7), 602–618. 10.1016/S1474-4422(23)00131-X

Elbaz, A., Vicente-Vytopilova, P., Tavernier, B., Sabia, S., Dumurgier, J., Mazoyer, B., Singh-Manoux, A., & Tzourio, C. (2013). Motor function in the elderly: Evidence for the reserve hypothesis. Neurology, 81(5), 417–426. 10.1212/WNL.0b013e31829d8761

Ferrucci, L., & Kuchel, G. A. (2021). Heterogeneity of Aging: Individual Risk Factors, Mechanisms, Patient Priorities, and Outcomes. Journal of the American Geriatrics Society, 69(3), 610–612. 10.1111/jgs.17011

Franzmeier, N., Caballero, M. Á. A., Taylor, A. N. W., Simon-Vermot, L., Buerger, K., Ertl-Wagner, B., Mueller, C., Catak, C., Janowitz, D., Baykara, E., Gesierich, B., Duering, M., Ewers, M., & for the Alzheimer’s Disease Neuroimaging Initiative. (2017). Resting-state global functional connectivity as a biomarker of cognitive reserve in mild cognitive impairment. Brain Imaging and Behavior, 11(2), 368–382. 10.1007/s11682-016-9599-1

Gluhm, S., Goldstein, J., Loc, K., Colt, A., Liew, C. V., & Corey-Bloom, J. (2013). Cognitive Performance on the Mini-Mental State Examination and the Montreal Cognitive Assessment Across the Healthy Adult Lifespan. Cognitive and Behavioral Neurology, 26(1), 1. 10.1097/WNN.0b013e31828b7d26

Groemping, U. (2005). relaimpo: Relative Importance of Regressors in Linear Models (p. 2.2–7) [Dataset]. 10.32614/CRAN.package.relaimpo

Grotz, C., Seron, X., Van Wissen, M., & Adam, S. (2017). How should proxies of cognitive reserve be evaluated in a population of healthy older adults? International Psychogeriatrics, 29(1), 123–136. 10.1017/S1041610216001745

Haut, M. W., Moran, M. T., Lancaster, M. A., Kuwabara, H., Parsons, M. W., & Puce, A. (2007). White Matter Correlates of Cognitive Capacity Studied With Diffusion Tensor Imaging: Implications for Cognitive Reserve. Brain Imaging and Behavior, 1(3), 83–92. 10.1007/s11682-007-9008-x

Henschel, L., Conjeti, S., Estrada, S., Diers, K., Fischl, B., & Reuter, M. (2020). FastSurfer— A fast and accurate deep learning based neuroimaging pipeline. NeuroImage, 219, 117012. 10.1016/j.neuroimage.2020.117012

Hoopes, A., Mora, J. S., Dalca, A. V., Fischl, B., & Hoffmann, M. (2022). SynthStrip: Skull-stripping for any brain image. NeuroImage, 260, 119474. 10.1016/j.neuroimage.2022.119474

Hua, K., Zhang, J., Wakana, S., Jiang, H., Li, X., Reich, D. S., Calabresi, P. A., Pekar, J. J., van Zijl, P. C. M., & Mori, S. (2008). Tract probability maps in stereotaxic spaces: Analyses of white matter anatomy and tract-specific quantification. NeuroImage, 39(1), 336–347. 10.1016/j.neuroimage.2007.07.053

Jenkinson, M., Bannister, P., Brady, M., & Smith, S. (2002). Improved optimization for the robust and accurate linear registration and motion correction of brain images. NeuroImage, 17(2), 825–841. 10.1016/s1053-8119(02)91132-8

Jin, Y., Lin, L., Xiong, M., Sun, S., & Wu, S.-C. (2023). Moderating effects of cognitive reserve on the relationship between brain structure and cognitive abilities in middle-aged and older adults. Neurobiology of Aging, 128, 49–64. 10.1016/j.neurobiolaging.2023.04.003

Jolly, T. A. D., Cooper, P. S., Wan Ahmadul Badwi, S. A., Phillips, N. A., Rennie, J. L., Levi, C. R., Drysdale, K. A., Parsons, M. W., Michie, P. T., & Karayanidis, F. (2016). Microstructural white matter changes mediate age-related cognitive decline on the Montreal Cognitive Assessment (MoCA). Psychophysiology, 53(2), 258–267. 10.1111/psyp.12565

Jones, R. N., Manly, J., Glymour, M. M., Rentz, D. M., Jefferson, A. L., & Stern, Y. (2011). Conceptual and measurement challenges in research on cognitive reserve. Journal of the International Neuropsychological Society: JINS, 17(4), 593–601. 10.1017/S1355617710001748

Katzman, R., Terry, R., DeTeresa, R., Brown, T., Davies, P., Fuld, P., Renbing, X., & Peck, A. (1988). Clinical, pathological, and neurochemical changes in dementia: A subgroup with preserved mental status and numerous neocortical plaques. Annals of Neurology, 23(2), 138–144. 10.1002/ana.410230206

Kerchner, G. A., Racine, C. A., Hale, S., Wilheim, R., Laluz, V., Miller, B. L., & Kramer, J. H. (2012). Cognitive Processing Speed in Older Adults: Relationship with White Matter Integrity. PLOS ONE, 7(11), e50425. 10.1371/journal.pone.0050425

Kobayashi-Cuya, K. E., Sakurai, R., Sakuma, N., Suzuki, H., Ogawa, S., Takebayashi, T., & Fujiwara, Y. (2024). Bidirectional associations of high-level cognitive domains with hand motor function and gait speed in high-functioning older adults: A 7-year study. Archives of Gerontology and Geriatrics, 117, 105232. 10.1016/j.archger.2023.105232

Kobayashi-Cuya, K. E., Sakurai, R., Suzuki, H., Ogawa, S., Takebayashi, T., & Fujiwara, Y. (2018). Observational Evidence of the Association Between Handgrip Strength, Hand Dexterity, and Cognitive Performance in Community-Dwelling Older Adults: A Systematic Review. Journal of Epidemiology, 28(9), 373–381. 10.2188/jea.JE20170041

Koçer, A., & Oktay, A. B. (2016). Nintendo Wii assessment of Hoehn and Yahr score with Parkinson’s disease tremor. Technology and Health Care: Official Journal of the European Society for Engineering and Medicine, 24(2), 185–191. 10.3233/THC-151124

Kong, T. S., Gratton, C., Low, K. A., Tan, C. H., Chiarelli, A. M., Fletcher, M. A., Zimmerman, B., Maclin, E. L., Sutton, B. P., Gratton, G., & Fabiani, M. (2020). Age-related differences in functional brain network segregation are consistent with a cascade of cerebrovascular, structural, and cognitive effects. Network Neuroscience, 4(1), 89–114. 10.1162/netn_a_00110

Lavrencic, L. M., Churches, O. F., & Keage, H. A. D. (2018). Cognitive reserve is not associated with improved performance in all cognitive domains. Applied Neuropsychology: Adult, 25(5), 473–485. 10.1080/23279095.2017.1329146

Law, D. J., Morrin, K. A., & Pellegrino, J. W. (1995). Training effects and working memory contributions to skill acquisition in a complex coordination task. Learning and Individual Differences, 7(3), 207–234. 10.1016/1041-6080(95)90011-X

Leisman, G., Moustafa, A. A., & Shafir, T. (2016). Thinking, Walking, Talking: Integratory Motor and Cognitive Brain Function. Frontiers in Public Health, 4. 10.3389/fpubh.2016.00094

Lin, L., Jin, Y., Xiong, M., Wu, S., & Sun, S. (2023). The Protective Power of Cognitive Reserve: Examining White Matter Integrity and Cognitive Function in the Aging Brain for Sustainable Cognitive Health. Sustainability, 15(14), Article 14. 10.3390/su151411336

Liu, Y., Cai, Z.-L., Xue, S., Zhou, X., & Wu, F. (2013). Proxies of cognitive reserve and their effects on neuropsychological performance in patients with mild cognitive impairment. Journal of Clinical Neuroscience: Official Journal of the Neurosurgical Society of Australasia, 20(4), 548–553. 10.1016/j.jocn.2012.04.020

Liu, Y., Julkunen, V., Paajanen, T., Westman, E., Wahlund, L.-O., Aitken, A., Sobow, T., Mecocci, P., Tsolaki, M., Vellas, B., Muehlboeck, S., Spenger, C., Lovestone, S., Simmons, A., Soininen, H., & AddNeuroMed Consortium. (2012). Education increases reserve against Alzheimer’s disease—Evidence from structural MRI analysis. Neuroradiology, 54(9), 929–938. 10.1007/s00234-012-1005-0

Livingston, G., Huntley, J., Liu, K. Y., Costafreda, S. G., Selbæk, G., Alladi, S., Ames, D., Banerjee, S., Burns, A., Brayne, C., Fox, N. C., Ferri, C. P., Gitlin, L. N., Howard, R., Kales, H. C., Kivimäki, M., Larson, E. B., Nakasujja, N., Rockwood, K.,… Mukadam, N. (2024). Dementia prevention, intervention, and care: 2024 report of the Lancet standing Commission. The Lancet, 404(10452), 572–628. 10.1016/S0140-6736(24)01296-0

Logan, G. D., & Cowan, W. B. (1984). On the ability to inhibit thought and action: A theory of an act of control. Psychological Review, 91(3), 295–327. 10.1037/0033-295X.91.3.295

Long, J. A. (2019). interactions: Comprehensive, User-Friendly Toolkit for Probing Interactions. CRAN: Contributed Packages. 10.32614/cran.package.interactions

Lu, C. H., & Proctor, R. W. (1995). The influence of irrelevant location information on performance: A review of the Simon and spatial Stroop effects. Psychonomic Bulletin & Review, 2(2), 174–207. 10.3758/BF03210959

Madden, D. J., Bennett, I. J., Burzynska, A., Potter, G. G., Chen, N., & Song, A. W. (2012). Diffusion tensor imaging of cerebral white matter integrity in cognitive aging. Biochimica et Biophysica Acta (BBA) - Molecular Basis of Disease, 1822(3), 386–400. 10.1016/j.bbadis.2011.08.003

Montemurro, S., Daini, R., Tagliabue, C., Guzzetti, S., Gualco, G., Mondini, S., & Arcara, G. (2023). Cognitive reserve estimated with a life experience questionnaire outperforms education in predicting performance on MoCA: Italian normative data. Current Psychology, 42(23), 19503–19517. 10.1007/s12144-022-03062-6

Montemurro, S., Filippini, N., Ferrazzi, G., Mantini, D., Arcara, G., & Marino, M. (2023). Education differentiates cognitive performance and resting state fMRI connectivity in healthy aging. Frontiers in Aging Neuroscience, 15, 1168576. 10.3389/fnagi.2023.1168576

Nasreddine, Z. S., Phillips, N. A., Bédirian, V., Charbonneau, S., Whitehead, V., Collin, I., Cummings, J. L., & Chertkow, H. (2005). The Montreal Cognitive Assessment, MoCA: A brief screening tool for mild cognitive impairment. Journal of the American Geriatrics Society, 53(4), 695–699. 10.1111/j.1532-5415.2005.53221.x

Nelson, M. E., Jester, D. J., Petkus, A. J., & Andel, R. (2021). Cognitive Reserve, Alzheimer’s Neuropathology, and Risk of Dementia: A Systematic Review and Meta-Analysis. Neuropsychology Review, 31(2), 233–250. 10.1007/s11065-021-09478-4

Nickerson, L. D., Smith, S. M., Öngür, D., & Beckmann, C. F. (2017). Using Dual Regression to Investigate Network Shape and Amplitude in Functional Connectivity Analyses. Frontiers in Neuroscience, 11, 115. 10.3389/fnins.2017.00115

Nogueira, J., Gerardo, B., Santana, I., Simões, M. R., & Freitas, S. (2022). The Assessment of Cognitive Reserve: A Systematic Review of the Most Used Quantitative Measurement Methods of Cognitive Reserve for Aging. Frontiers in Psychology, 13. https://www.frontiersin.org/articles/10.3389/fpsyg.2022.847186

Nucci, M., Mapelli, D., & Mondini, S. (2012). Cognitive Reserve Index questionnaire (CRIq): A new instrument for measuring cognitive reserve. Aging Clinical and Experimental Research, 24(3), 218–226. 10.3275/7800

Nyberg, L., Lövdén, M., Riklund, K., Lindenberger, U., & Bäckman, L. (2012). Memory aging and brain maintenance. Trends in Cognitive Sciences, 16(5), 292–305. 10.1016/j.tics.2012.04.005

Opdebeeck, C., Martyr, A., & Clare, L. (2016). Cognitive reserve and cognitive function in healthy older people: A meta-analysis. Aging, Neuropsychology, and Cognition, 23(1), 40–60. 10.1080/13825585.2015.1041450

Panico, F., Sagliano, L., Magliacano, A., Santangelo, G., & Trojano, L. (2022). The relationship between cognitive reserve and cognition in healthy adults: A systematic review. Current Psychology. 10.1007/s12144-022-03523-y

Pedregosa, F., Varoquaux, G., Gramfort, A., Michel, V., Thirion, B., Grisel, O., Blondel, M., Prettenhofer, P., Weiss, R., Dubourg, V., Vanderplas, J., Passos, A., Cournapeau, D., Brucher, M., Perrot, M., & Duchesnay, É. (2011). Scikit-learn: Machine Learning in Python. J. Mach. Learn. Res., 12(null), 2825–2830.

Poldrack, R. A., Mumford, J. A., & Nichols, T. E. (2011). Handbook of Functional MRI Data Analysis. Cambridge University Press. 10.1017/CBO9780511895029

Querbes, O., Aubry, F., Pariente, J., Lotterie, J.-A., Démonet, J.-F., Duret, V., Puel, M., Berry, I., Fort, J.-C., Celsis, P., & The Alzheimer’s Disease Neuroimaging Initiative. (2009). Early diagnosis of Alzheimer’s disease using cortical thickness: Impact of cognitive reserve. Brain, 132(8), 2036–2047. 10.1093/brain/awp105

Quinzi, F., Berchicci, M., Bianco, V., Di Filippo, G., Perri, R. L., & Di Russo, F. (2020). The role of cognitive reserve on prefrontal and premotor cortical activity in visuo-motor response tasks in healthy old adults. Neurobiology of Aging, 94, 185–195. 10.1016/j.neurobiolaging.2020.06.002

R Core Team. (2022). R: A language and environment for statistical computing. R Foundation for Statistical Computing, Vienna, Austria. [Computer software]. https://www.R-project.org/

Ratcliff, R., & Smith, P. L. (2004). A Comparison of Sequential Sampling Models for Two-Choice Reaction Time. Psychological Review, 111(2), 333–367. 10.1037/0033-295X.111.2.333

Raven, J., & Raven, J. (2003). Raven Progressive Matrices. In Handbook of nonverbal assessment (pp. 223–237). Kluwer Academic/Plenum Publishers. 10.1007/978-1-4615-0153-4_11

Raz, N., Lindenberger, U., Rodrigue, K. M., Kennedy, K. M., Head, D., Williamson, A., Dahle, C., Gerstorf, D., & Acker, J. D. (2005). Regional Brain Changes in Aging Healthy Adults: General Trends, Individual Differences and Modifiers. Cerebral Cortex, 15(11), 1676–1689. 10.1093/cercor/bhi044

Rentz, D. M., Locascio, J. J., Becker, J. A., Moran, E. K., Eng, E., Buckner, R. L., Sperling, R. A., & Johnson, K. A. (2010). Cognition, reserve, and amyloid deposition in normal aging. Annals of Neurology, 67(3), 353–364. 10.1002/ana.21904

Ritchie, S. J., Bates, T. C., Der, G., Starr, J. M., & Deary, I. J. (2013). Education is associated with higher later life IQ scores, but not with faster cognitive processing speed. Psychology and Aging, 28(2), 515–521. 10.1037/a0030820

Salimi-Khorshidi, G., Douaud, G., Beckmann, C. F., Glasser, M. F., Griffanti, L., & Smith, S. M. (2014). Automatic denoising of functional MRI data: Combining independent component analysis and hierarchical fusion of classifiers. NeuroImage, 90, 449–468. 10.1016/j.neuroimage.2013.11.046

Salthouse, T. A. (2009). When does age-related cognitive decline begin? Neurobiology of Aging, 30(4), 507–514. 10.1016/j.neurobiolaging.2008.09.023

Salthouse, T. A. (2011). Neuroanatomical substrates of age-related cognitive decline. Psychological Bulletin, 137(5), 753–784. 10.1037/a0023262

Sánchez-Izquierdo, M., & Fernández-Ballesteros, R. (2021). Cognition in Healthy Aging. International Journal of Environmental Research and Public Health, 18(3), Article 3. 10.3390/ijerph18030962

Saywell, I., Foreman, L., Child, B., Phillips-Hughes, A. L., Collins-Praino, L., & Baetu, I. (2024). Influence of cognitive reserve on cognitive and motor function in α-synucleinopathies: A systematic review and multilevel meta-analysis. Neuroscience & Biobehavioral Reviews, 161, 105672. 10.1016/j.neubiorev.2024.105672

Schaie, K. W., & Willis, S. L. (2010). The Seattle Longitudinal Study of Adult Cognitive Development. ISSBD Bulletin, 57(1), 24–29.

Seidler, R. D., Bernard, J. A., Burutolu, T. B., Fling, B. W., Gordon, M. T., Gwin, J. T., Kwak, Y., & Lipps, D. B. (2010). Motor Control and Aging: Links to Age-Related Brain Structural, Functional, and Biochemical Effects. Neuroscience and Biobehavioral Reviews, 34(5), 721–733. 10.1016/j.neubiorev.2009.10.005

Smith, S. M., Jenkinson, M., Johansen-Berg, H., Rueckert, D., Nichols, T. E., Mackay, C. E., Watkins, K. E., Ciccarelli, O., Cader, M. Z., Matthews, P. M., & Behrens, T. E. J. (2006). Tract-based spatial statistics: Voxelwise analysis of multi-subject diffusion data. NeuroImage, 31(4), 1487–1505. 10.1016/j.neuroimage.2006.02.024

Stern, Y. (2002). What is cognitive reserve? Theory and research application of the reserve concept. Journal of the International Neuropsychological Society, 8(3), 448–460. 10.1017/S1355617702813248

Stern, Y. (2009). Cognitive reserve. Neuropsychologia, 47(10), 2015–2028. 10.1016/j.neuropsychologia.2009.03.004

Stern, Y., Albert, M., Barnes, C. A., Cabeza, R., Pascual-Leone, A., & Rapp, P. R. (2023). A framework for concepts of reserve and resilience in aging. Neurobiology of Aging, 124, 100–103. 10.1016/j.neurobiolaging.2022.10.015

Stern, Y., Gazes, Y., Razlighi, Q., Steffener, J., & Habeck, C. (2018). A task-invariant cognitive reserve network. NeuroImage, 178, 36–45. 10.1016/j.neuroimage.2018.05.033

Tucker, A. M., & Stern, Y. (2011). Cognitive reserve in aging. Current Alzheimer Research, 8(4), 354–360. 10.2174/156720511795745320

Tucker-Drob, E. M., Johnson, K. E., & Jones, R. N. (2009). The cognitive reserve hypothesis: A longitudinal examination of age-associated declines in reasoning and processing speed. Developmental Psychology, 45, 431–446. 10.1037/a0014012

Valenzuela, M. J., & Sachdev, P. (2006a). Brain reserve and cognitive decline: A non-parametric systematic review. Psychological Medicine, 36(8), 1065–1073. 10.1017/S0033291706007744

Valenzuela, M. J., & Sachdev, P. (2006b). Brain reserve and dementia: A systematic review. Psychological Medicine, 36(4), 441–454. 10.1017/S0033291705006264

Valenzuela, M. J., Sachdev, P., Wen, W., Chen, X., & Brodaty, H. (2008). Lifespan Mental Activity Predicts Diminished Rate of Hippocampal Atrophy. PLOS ONE, 3(7), e2598. 10.1371/journal.pone.0002598

Varela-López, B., Cruz-Gómez, Á. J., Lojo-Seoane, C., Díaz, F., Pereiro, A. X., Zurrón, M., Lindín, M., & Galdo-Álvarez, S. (2022). Cognitive reserve, neurocognitive performance, and high-order resting-state networks in cognitively unimpaired aging. Neurobiology of Aging, 117, 151–164. 10.1016/j.neurobiolaging.2022.05.012

Vickers, D., Nettelbeck, T., & Willson, R. J. (1972). Perceptual Indices of Performance: The Measurement of ‘Inspection Time’ and ‘Noise’ in the Visual System. Perception, 1(3), 263–295. 10.1068/p010263

Viechtbauer, W. (2010). Conducting Meta-Analyses in R with the metafor Package. Journal of Statistical Software, 36, 1–48. 10.18637/jss.v036.i03

Vuoksimaa, E., Panizzon, M. S., Chen, C.-H., Eyler, L. T., Fennema-Notestine, C., Fiecas, M. J. A., Fischl, B., Franz, C. E., Grant, M. D., Jak, A. J., Lyons, M. J., Neale, M. C., Thompson, W. K., Tsuang, M. T., Xian, H., Dale, A. M., & Kremen, W. S. (2013). Cognitive reserve moderates the association between hippocampal volume and episodic memory in middle age. Neuropsychologia, 51(6), 1124–1131. 10.1016/j.neuropsychologia.2013.02.022

Wei, N., Deng, Y., Yao, L., Jia, W., Wang, J., Shi, Q., Chen, H., Pan, Y., Yan, H., Zhang, Y., & Wang, Y. (2019). A Neuroimaging Marker Based on Diffusion Tensor Imaging and Cognitive Impairment Due to Cerebral White Matter Lesions. Frontiers in Neurology, 10. 10.3389/fneur.2019.00081

Weiler, M., Casseb, R. F., de Campos, B. M., de Ligo Teixeira, C. V., Carletti-Cassani, A. F. M. K., Vicentini, J. E., Magalhães, T. N. C., de Almeira, D. Q., Talib, L. L., Forlenza, O. V., Balthazar, M. L. F., & Castellano, G. (2018). Cognitive Reserve Relates to Functional Network Efficiency in Alzheimer’s Disease. Frontiers in Aging Neuroscience, 10, 255. 10.3389/fnagi.2018.00255

Wilson, R. S., Beckett, L. A., Barnes, L. L., Schneider, J. A., Bach, J., Evans, D. A., & Bennett, D. A. (2002). Individual differences in rates of change in cognitive abilities of older persons. Psychology and Aging, 17(2), 179–193. 10.1037/0882-7974.17.2.179

Woods, D. L., Kishiyama, M. M., Yund, E. W., Herron, T. J., Edwards, B., Poliva, O., Hink, R. F., & Reed, B. (2011). Improving digit span assessment of short-term verbal memory. Journal of Clinical and Experimental Neuropsychology, 33(1), 101–111. 10.1080/13803395.2010.493149

Yang, W., Wang, J., Guo, J., Dove, A., Qi, X., Bennett, D. A., & Xu, W. (2024). Association of Cognitive Reserve Indicator with Cognitive Decline and Structural Brain Differences in Middle and Older Age: Findings from the UK Biobank. The Journal of Prevention of Alzheimer’s Disease, 11(3), 739–748. 10.14283/jpad.2024.54

Yang, Y., Chen, Y., Yang, C., Chen, K., Li, X., & Zhang, Z. (2022). Contributions of early-life cognitive reserve and late-life leisure activity to successful and pathological cognitive aging. BMC Geriatrics, 22(1), 831. 10.1186/s12877-022-03530-5

Ye, Q., Zhu, H., Chen, H., Liu, R., Huang, L., Chen, H., Cheng, Y., Qin, R., Shao, P., Xu, H., Ma, J., & Xu, Y. (2022). Effects of cognitive reserve proxies on cognitive function and frontoparietal control network in subjects with white matter hyperintensities: A cross-sectional functional magnetic resonance imaging study. CNS Neuroscience & Therapeutics, 28(6), 932–941. 10.1111/cns.13824

