## Supplementary Material for "Lifetime experiences as proxies of cognitive reserve predict cognition and motor function beyond multimodal MRI brain measures in healthy adults"

**Table 1**

Degree of imputation used for specific cognitive and motor function tests prior to principal component analyses

| Test | Imputation count |
|---|---|
| Cognition measures | |
| Raven's matrices | 1 |
| Inspection time | 3 |
| Two-choice decision time | 5 |
| Dot matrix | 0 |
| Stop signal task | 3 |
| Simon task | 0 |
| Digit span total | 1 |
| MoCA | 0 |
| Motor function measures | |
| Tapping test | 0 |
| Two-choice response time | 5 |
| Tremor (theta power) | 0 |
| Balance (path length feet together) | 1 |

Imputation was performed using the median value.

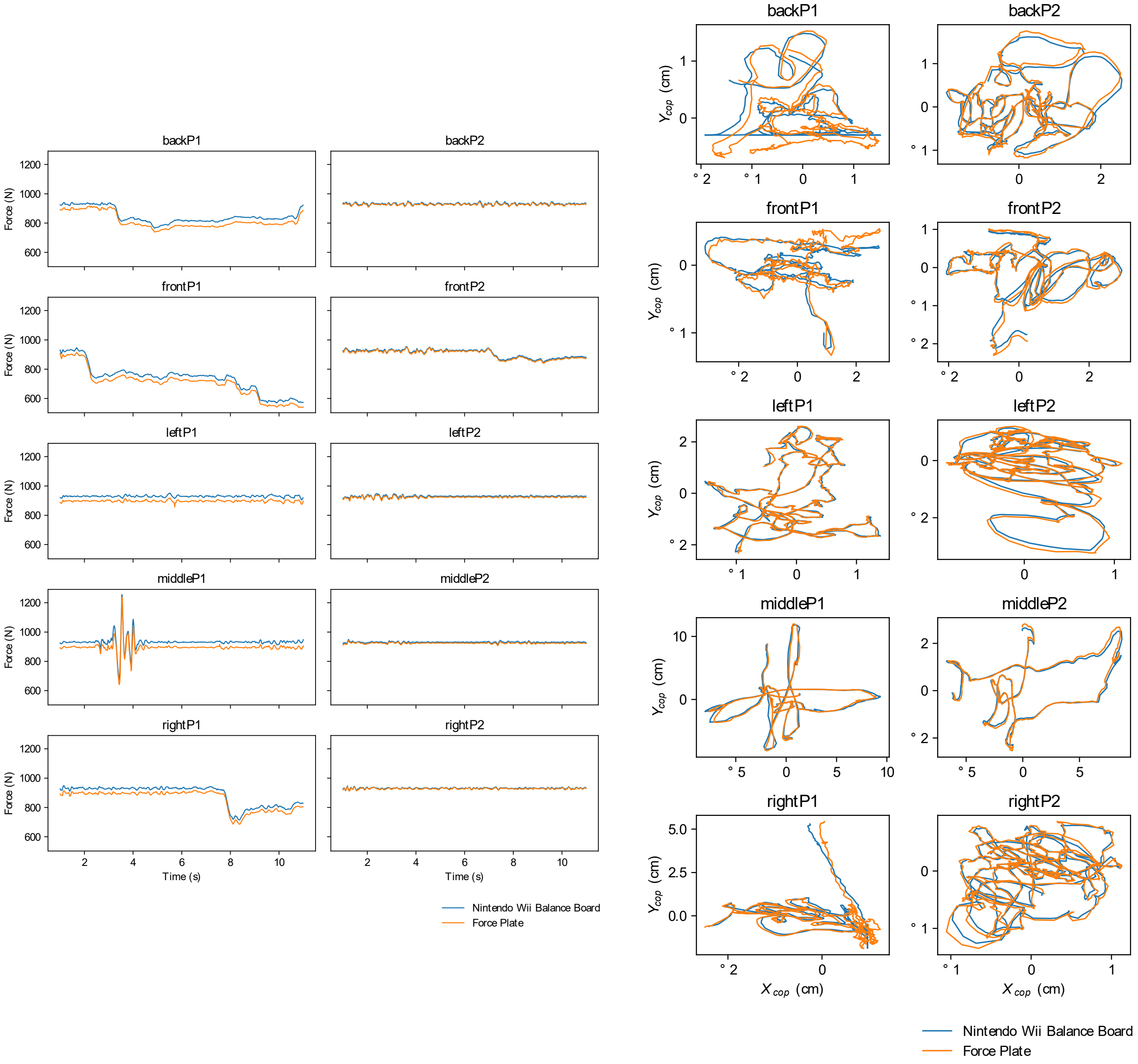

**Fig 1.** Comparison of raw time series (left) and centre of pressure path length (right) balance data simultaneously obtained from a laboratory grade force platform (orange) and a Nintendo® Wii balance board used in the current study (blue). Recordings were taken whilst a volunteer stood positioned on either the back, front, left, or right of both devices simultaneously; two 30-second recordings were completed at each position.

**Table 2**

Mapping of FastSurfer structural segmentations into a final list of brain regions used to create a composite structural brain measure for analyses.

| **FastSurfer Segmentations** | **Merged Brain Region** | **Final Structural Brain Measure List** |
|---|---|---|
| Cerebral WM | | Cerebral WM volume |
| Cerebellum GM, Cerebellum WM | Cerebellum | Cerebellum volume |
| Thalamus | | Thalamus volume |
| Putamen, Caudate | Striatum | Striatum volume |
| Globus Pallidus | | Globus Pallidus volume |
| Nucleus Accumbens | | Nucleus Accumbens volume |
| Hippocampus | | Hippocampus volume |
| Amygdala | | Amygdala volume |
| Ventral Diencephalon | | Ventral Diencephalon volume |
| Brainstem | | Brainstem volume |
| Rostral ACC, Caudal ACC, PCC, Isthmus of the Cingulate | Cingulate Cortex | Cingulate Cortex volume |
| Anterior CC, Mid-Anterior CC, Central CC, Mid-Posterior CC, Posterior CC | Corpus Callosum | Corpus Callosum volume |
| Lateral Ventricle, Inferior Lateral Ventricle, Third Ventricle, Fourth Ventricle | Ventricular CSF | Ventricular CSF volume |
| Pars Opercularis, Pars Triangularis, Pars Orbitalis | Inferior Frontal Gyrus | Inferior Frontal Gyrus volume |
| Lateral Orbitofrontal Cortex, Medial Orbitofrontal Cortex | Orbitofrontal Cortex | Orbitofrontal Cortex volume |
| Rostral Middle Frontal Gyrus, Caudal Middle Frontal Gyrus | Middle Frontal Gyrus | Middle Frontal Gyrus volume |
| Superior Frontal Gyrus | | Superior Frontal Gyrus volume |

| | | |
|---|---|---|
| Precentral Gyrus | | Precentral Gyrus volume |
| Postcentral Gyrus | | Postcentral Gyrus volume |
| Paracentral Gyrus | | Paracentral Gyrus volume |
| Inferior Parietal Gyrus, Superior Parietal Gyrus | Parietal Lobule | Parietal Lobule volume |
| Supramarginal Gyrus | | Supramarginal Gyrus volume |
| Precuneus | | Precuneus volume |
| Insula | | Insula volume |
| Inferior Temporal Gyrus, Middle Temporal Gyrus, Superior Temporal Gyrus, Transverse Temporal Gyrus | Temporal Gyrus | Temporal Gyri volume |
| Parahippocampal Gyrus | | Parahippocampal Gyrus volume |
| Fusiform Gyrus | | Fusiform Gyrus volume |
| Entorhinal Cortex | | Entorhinal Cortex volume |
| Cuneus, Lingual, Lateral Occipital Gyrus, Pericalcarine Gyrus | Occipital Lobe | Occipital Lobe volume |
| Rostral ACC, Caudal ACC, Pars Opercularis, Pars Triangularis, Pars Orbitalis, Lateral Orbitofrontal Cortex, Medial Orbitofrontal Cortex, Rostral Middle Frontal Gyrus, Caudal Middle Frontal Gyrus, Superior Frontal Gyrus, Precentral Gyrus | Frontal Lobe | Frontal Lobe thickness |
| Postcentral Gyrus, Inferior Parietal Gyrus, Superior Parietal Gyrus, Supramarginal Gyrus, Precuneus | Parietal Lobe | Parietal Lobe thickness |
| Insula, Inferior Temporal Gyrus, Middle Temporal Gyrus, Superior Temporal Gyrus, Transverse Temporal Gyrus, Parahippocampal Gyrus, Fusiform Gyrus, Entorhinal Cortex | Temporal Lobe | Temporal Lobe thickness |
| Cuneus, Lingual, Lateral Occipital Gyrus, Pericalcarine Gyrus | Occipital Lobe | Occipital Lobe thickness |

FastSurfer segmentations grouped together in the ‘FastSurfer Segmentations’ column were merged into a single region identified in the ‘Merged Brain Region’ column. Combined brain regions were summed together, as well as left and right structures, if they are present bilaterally. ACC = anterior cingulate cortex. CC = corpus callosum. CSF = cerebrospinal fluid. GM = grey matter. PCC = posterior cingulate cortex. WM = white matter.

**Table 3**

Mapping of John Hopkins University International Consortium for Brain Mapping white matter tract atlas into a final list of tracts used to create a composite diffusion brain measure for analyses.

| JHU-ICBM WM Tracts Atlas | Merged Tracts | Final Diffusion Brain Measure List |
|---|---|---|
| Middle Cerebellar Peduncle, Pontine Crossing Tract, Inferior Cerebellar Peduncle, Superior Cerebellar Peduncle | Cerebellar Peduncle | Cerebellar Peduncle FA |
| Genu of CC, Body of CC, Splenium of CC | CC | CC FA |
| Fornix (body and column) | | Fornix FA |
| Corticospinal Tract | | Corticospinal Tract FA |
| Medial Lemniscus | | Medial Lemniscus FA |
| Cerebral Peduncle | | Cerebral Peduncle FA |
| Anterior Limb of Internal Capsule, Posterior Limb of Internal Capsule, Retrolenticular Limb of Internal Capsule | Internal Capsule | Internal Capsule FA |
| Anterior Corona Radiata. Superior Corona Radiata, Posterior Corona Radiata | Corona Radiata | Corona Radiata FA |
| Posterior Thalamic Radiation | | Posterior Thalamic Radiation FA |
| Sagittal Stratum | | Sagittal Stratum FA |
| External Capsule | | External Capsule FA |
| Cingulum (cingulate gyrus), Cingulum (hippocampus) | Cingulum | Cingulum FA |
| Stria Terminalis | | Stria Terminalis FA |
| Superior Longitudinal Fasciculus | | Superior Longitudinal Fasciculus FA |

| | | |
|---|---|---|
| Superior Fronto-occipital Fasciculus, Inferior Fronto-occipital Fasciculus | Fronto-occipital Fasciculus | Fronto-occipital Fasciculus FA |
| Uncinate Fasciculus | | Uncinate Fasciculus FA |
| Tapetum | | Tapetum FA |

Tracts from the John Hopkins University International Consortium for Brain Mapping atlas were grouped together in the 'JHU-ICBM Atlas Tracts' column were merged into a single tract identified in the 'Merged Tracts' column. Combined white matter tracts were summed together, as well as left and right tracts, if they are present bilaterally. CC = corpus callosum. FA = fractional anisotropy. JHU-ICBM = John Hopkins University International Consortium for Brain Mapping. WM = white matter.

A

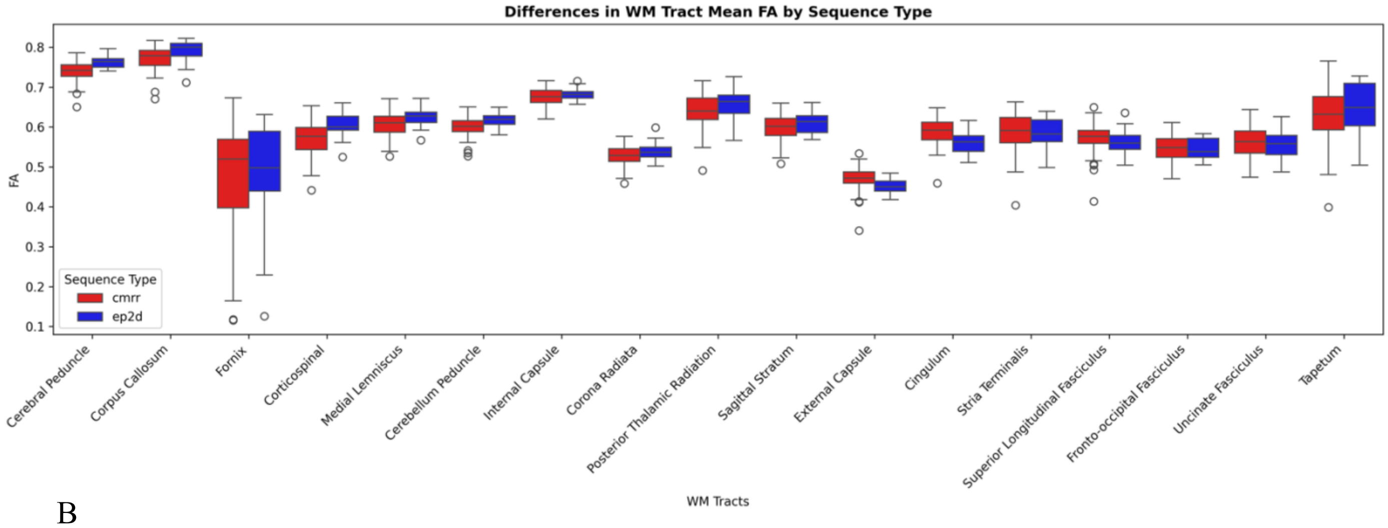

B

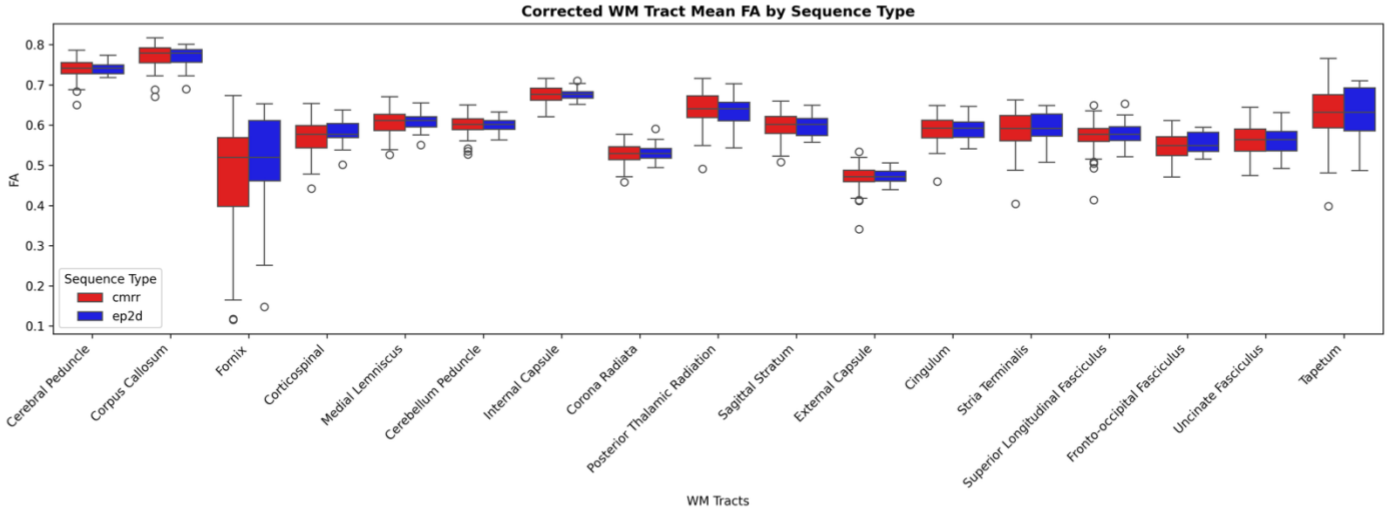

**Fig 2.** Boxplots demonstrating the differences in fractional anisotropy between each sequence type for white matter tracts (A) and the result of correcting data collected using the two-dimensional echo planar imaging diffusion sequence to the Center for Magnetic Resonance Research diffusion sequence (B). These differences were evaluated using the entire study sample (n=101) and neither sequence was exclusive to Skyra or Cima.X machines. CMRR = Center for Magnetic Resonance Research. EP2D = two-dimensional echo-planar imaging. FA = fractional anisotropy. WM = white matter.

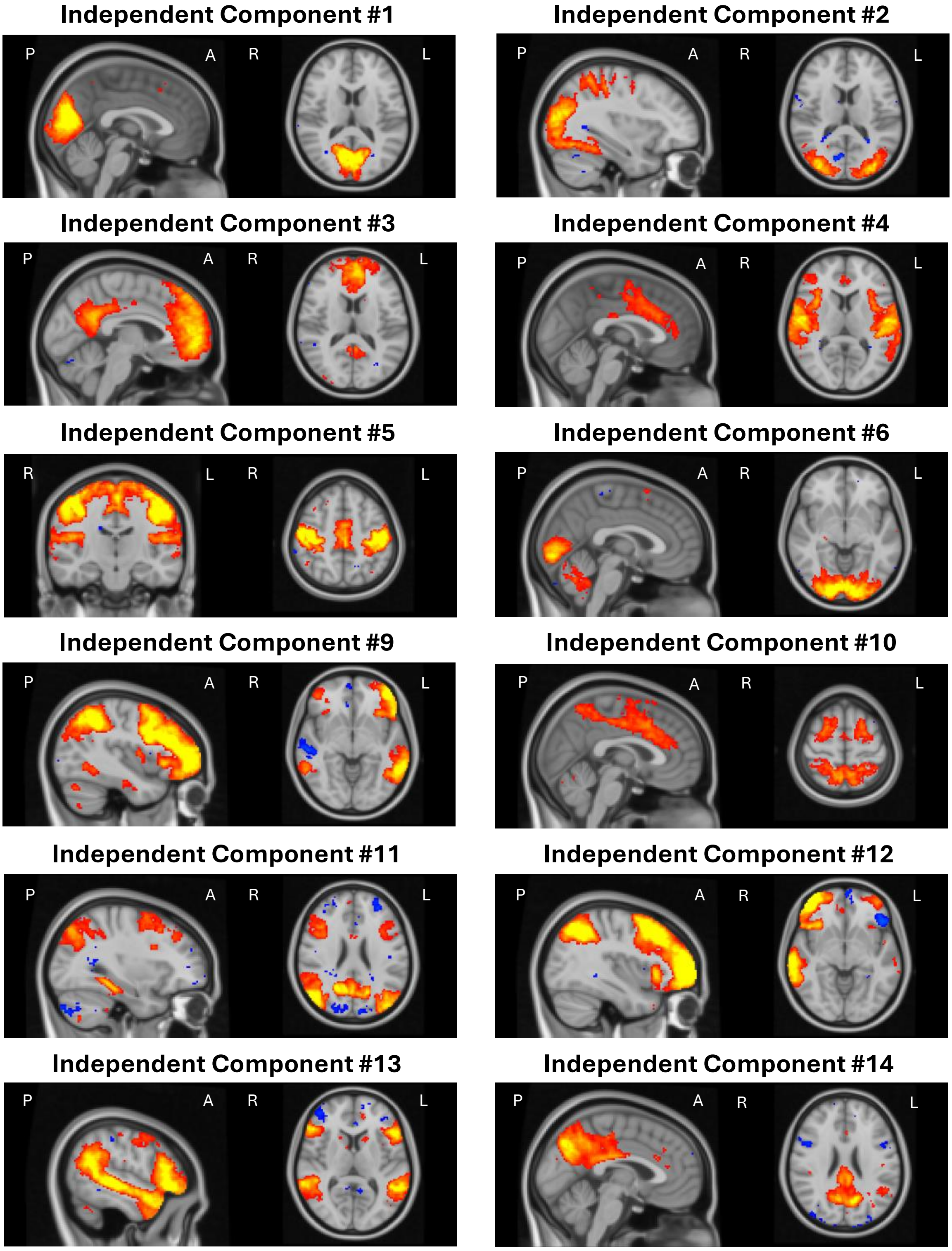

**Fig 3.** Group-level spatial maps of resting-state networks produced from group independent component analysis with dimensionality 20. Positive intensities (activity of interest) are highlighted in red-yellow. Brighter, more yellow colours represent greater positive intensity values (more activity). Resting-state network spatial maps have been overlaid on the Montreal Neurological Institute's 152 2mm brain template. Independent components classified as noise have not been displayed. A = anterior. L = left. P = posterior. R = right.

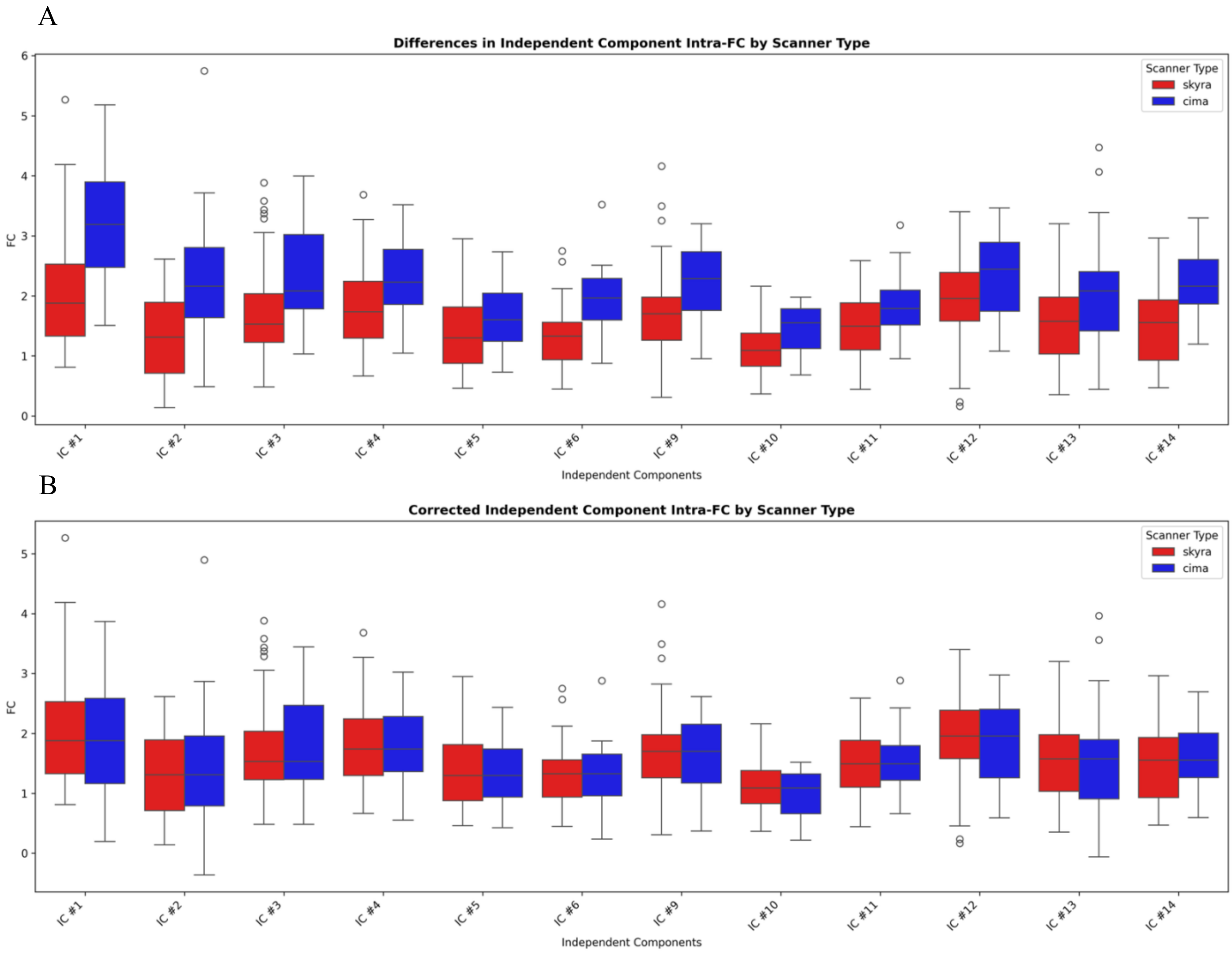

**Fig 4.** Boxplots demonstrating the differences in functional connectivity between each scanner type for neural signal independent components (A) and the result of correcting data collected using the Cima.X machine to the Skyra machine (B). These differences were evaluated using the entire study sample (n=101). FC = functional connectivity. IC = independent component.

**Table 4**

Summary statistics for each MRI brain measure included in principal component analyses.

| Structural | | Diffusion | | Functional | |
|---|---|---|---|---|---|
| Measure | M ± SD | Measure | M ± SD | Measure | M ± SD |
| Cerebral WM volume | 467914.01 ± 60905.24 | Cerebellum Peduncles FA | 0.60 ± 0.02 | Independent Component #1 FC | 2.01 ± 0.96 |
| Cerebellum volume | 140266.66 ± 14167.14 | CC FA | 0.77 ± 0.03 | Independent Component #2 FC | 1.36 ± 0.79 |
| Thalamus volume | 15421.35 ± 1927.00 | Fornix FA | 0.48 ± 0.14 | Independent Component #3 FC | 1.73 ± 0.77 |
| Striatum volume | 16900.87 ± 1790.80 | Corticospinal Tract FA | 0.57 ± 0.04 | Independent Component #4 FC | 1.79 ± 0.65 |
| Globus Pallidus volume | 4218.96 ± 437.59 | Medial Lemniscus FA | 0.61 ± 0.03 | Independent Component #5 FC | 1.40 ± 0.62 |
| Nucleus Accumbens volume | 1085.33 ± 141.33 | Cerebral Peduncles FA | 0.74 ± 0.02 | Independent Component #6 FC | 1.29 ± 0.48 |
| Hippocampus volume | 8832.82 ± 754.09 | Internal Capsule FA | 0.68 ± 0.02 | Independent Component #9 FC | 1.72 ± 0.65 |
| Amygdala volume | 3595.08 ± 438.73 | Corona Radiata FA | 0.53 ± 0.03 | Independent Component #10 FC | 1.10 ± 0.39 |
| Ventral Diencephalon volume | 8638.55 ± 902.22 | Posterior Thalamic Radiation FA | 0.64 ± 0.04 | Independent Component #11 FC | 1.50 ± 0.52 |
| Brainstem volume | 21679.60 ± 2446.01 | Sagittal Stratum FA | 0.60 ± 0.03 | Independent Component #12 FC | 1.91 ± 0.67 |
| Cingulate Cortex volume | 22800.89 ± 2931.90 | External Capsule FA | 0.47 ± 0.03 | Independent Component #13 FC | 1.56 ± 0.75 |
| Corpus Callosum volume | 4304.59 ± 578.90 | Cingulum FA | 0.59 ± 0.03 | Independent Component #14 FC | 1.53 ± 0.61 |
| Ventricular CSF volume | 26644.34 ± 19686.30 | Stria Terminalis FA | 0.59 ± 0.05 | | |
| Inferior Frontal Gyrus volume | 12682.99 ± 1820.22 | Superior Longitudinal Fasciculus FA | 0.57 ± 0.03 | | |
| Orbitofrontal Cortex volume | 25968.94 ± 2962.28 | Frontal Occipital Fasciculus FA | 0.55 ± 0.03 | | |
| Middle Frontal Gyrus volume | 33225.86 ± 5131.14 | Uncinate Fasciculus FA | 0.56 ± 0.04 | | |
| Superior Frontal Gyrus volume | 48078.75 ± 5590.72 | Tapetum FA | 0.63 ± 0.07 | | |
| Precentral Gyrus volume | 25015.71 ± 2769.69 | | | | |

| | |
|---|---|
| Postcentral Gyrus volume | 20430.66 ± 2418.50 |
| Paracentral Gyrus volume | 8144.54 ± 941.75 |
| Parietal Lobule volume | 45963.43 ± 5564.69 |
| Supramarginal Gyrus volume | 19220.50 ± 2887.41 |
| Precuneus volume | 19309.63 ± 2862.91 |
| Insula volume | 12186.83 ± 1267.53 |
| Temporal Gyri volume | 84850.28 ± 9597.41 |
| Parahippocampal Gyrus volume | 4475.80 ± 454.49 |
| Fusiform Gyrus volume | 16732.41 ± 2279.97 |
| Entorhinal Cortex volume | 3830.51 ± 481.68 |
| Occipital Lobe volume | 49640.62 ± 6695.50 |
| Frontal Lobe thickness | 2.49 ± 0.09 |
| Parietal Lobe thickness | 2.36 ± 0.11 |
| Temporal Lobe thickness | 2.83 ± 0.13 |
| Occipital Lobe thickness | 1.89 ± 0.10 |

Summary statistics for each structural and diffusion brain measure used in principal component analyses have been reported, not original segmentation regions before combining structures or tracts. Raw data has been reported for structural MRI brain measures (not normalised by estimated total intracranial volume) in cubic millimetres ($mm^3$) (excluding thickness measures which are represented in millimetres). Bilateral brain regions and white matter tracts have been combined. CC = corpus callosum. CSF = cerebrospinal fluid. FA = fractional anisotropy. FC = functional connectivity. M = mean. SD = standard deviation. WM = white matter.

**Table 5**
Correlation matrix of raw data (significant correlation *p*-values ≤ .05 are shown in bold).

| Measure | 1 | 2 | 3 | 4 | 5 | 6 | 7 | 8 | 9 | 10 |
|---|---|---|---|---|---|---|---|---|---|---|
| 1. Age | | | | | | | | | | |
| 2. Sex | .048 | | | | | | | | | |
| 3. MoCA | -.146 | -.179 | | | | | | | | |
| 4. Education years | -.049 | .036 | .001 | | | | | | | |
| 5. Brain structure MRI composite | **-.539** | **-.439** | **.273** | .064 | | | | | | |
| 6. Brain diffusion MRI composite | **-.248** | **-.317** | **.325** | -.111 | **.468** | | | | | |
| 7. Brain functional MRI composite | **-.372** | -.135 | .048 | .15 | .115 | .145 | | | | |
| 8. CR proxy | **.787** | .015 | .146 | .095 | **-.374** | -.149 | **-.264** | | | |
| 9. Overall cognition composite | **-.609** | .158 | **.307** | .024 | **.294** | **.224** | **.206** | **-.352** | | |
| 10. Overall motor function composite | **-.398** | -.041 | **.351** | .099 | **.299** | **.214** | **.262** | -.141 | **.505** | |

CR = cognitive reserve. MoCA = Montreal Cognitive Assessment. MRI = magnetic resonance imaging.

**Table 6**

Correlation matrix of residuals generated for regression analyses (significant correlations *p*-values ≤ .05 are shown in bold).

| Measure | 1 | 2 | 3 | 4 | 5 | 6 | 7 | 8 |
|---|---|---|---|---|---|---|---|---|
| 1. MoCA | | | | | | | | |
| 2. Education years | .001 | | | | | | | |
| 3. Brain structure MRI composite | .145 | .083 | | | | | | |
| 4. Brain diffusion MRI composite | **.266** | -.121 | **.286** | | | | | |
| 5. Brain functional MRI composite | -.029 | .147 | **-.199** | .02 | | | | |
| 6. CR proxy | **.397** | **.248** | .077 | .049 | .033 | | | |
| 7. Overall cognition composite | **.345** | -.002 | .076 | .183 | -.003 | **.264** | | |
| 8. Overall motor function composite | **.322** | .085 | .099 | .132 | .132 | **.322** | **.34** | |

CR = cognitive reserve. MoCA = Montreal Cognitive Assessment. MRI = magnetic resonance imaging.

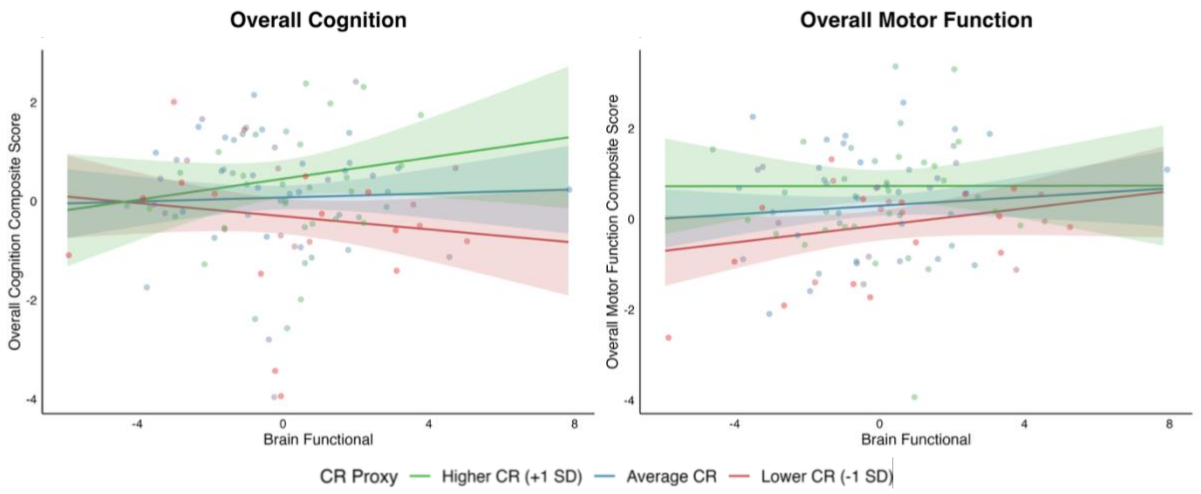

**Fig 5.** Interactions between cognitive reserve proxies and brain measure functional composite for overall cognition and overall motor function. CR = cognitive reserve.

**Table 7**

Regressions for specific cognitive tests (uncorrected *p*-values ≤ .05 are shown in bold).

| | Raven's matrices score | | | | Inspection time | | | | Two-choice decision time | | | | Dot matrix score | | | |
|---|---|---|---|---|---|---|---|---|---|---|---|---|---|---|---|---|
| | β | SE | *p* | $R^2$ | β | SE | *p* | $R^2$ | β | SE | *p* | $R^2$ | β | SE | *p* | $R^2$ |
| CR | | | | | | | | | | | | | | | | |
| CR proxy | 0.703 | 0.232 | **.003** | 0.082 | 4.979 | 2.455 | **.045** | 0.040 | 4.412 | 4.541 | .334 | 0.011 | 0.622 | 1.354 | .647 | 0.002 |
| MRI brain measures | | | | | | | | | | | | | | | | |
| Brain Structure | 0.056 | 0.070 | .430 | 0.017 | -0.860 | 0.744 | .250 | 0.008 | 0.093 | 1.376 | .946 | 0.002 | 0.162 | 0.410 | .693 | 0.001 |
| Brain Diffusion | 0.073 | 0.074 | .322 | 0.012 | 1.431 | 0.779 | .069 | 0.030 | 3.505 | 1.440 | **.017** | 0.061 | -0.462 | 0.429 | .284 | 0.011 |
| Brain Functional | -0.190 | 0.089 | **.035** | 0.042 | 0.458 | 0.939 | .627 | 0.005 | 3.632 | 1.737 | **.039** | 0.044 | 0.123 | 0.518 | .812 | 0.000 |
| Total Model $R^2$ | 0.153 (*p* = **.003**) | | | | 0.082 (*p* = .080) | | | | 0.117 (*p* = **.017**) | | | | 0.014 (*p* = .835) | | | |

| | Stop signal reaction time | | | | Simon task score | | | | Digit span total score | | | | MoCA score | | | |
|---|---|---|---|---|---|---|---|---|---|---|---|---|---|---|---|---|
| | β | SE | *p* | $R^2$ | β | SE | *p* | $R^2$ | β | SE | *p* | $R^2$ | β | SE | *p* | $R^2$ |
| CR | | | | | | | | | | | | | | | | |
| CR proxy | 8.663 | 6.586 | .192 | 0.017 | -4.720 | 4.478 | .294 | 0.011 | 0.678 | 0.369 | .069 | 0.035 | 0.770 | 0.182 | **<.001** | 0.151 |
| MRI brain measures | | | | | | | | | | | | | | | | |
| Brain Structure | -1.236 | 1.995 | .537 | 0.001 | 0.110 | 1.357 | .935 | 0.001 | 0.027 | 0.112 | .810 | 0.002 | 0.023 | 0.055 | .677 | 0.010 |
| Brain Diffusion | 3.524 | 2.089 | .095 | 0.027 | -1.450 | 1.420 | .310 | 0.011 | 0.093 | 0.117 | .430 | 0.008 | 0.144 | 0.058 | **.014** | 0.060 |
| Brain Functional | -2.467 | 2.519 | .330 | 0.007 | 1.417 | 1.713 | .410 | 0.006 | -0.016 | 0.141 | .911 | 0.000 | -0.029 | 0.069 | .680 | 0.001 |
| Total Model $R^2$ | 0.052 (*p* = .265) | | | | 0.029 (*p* = .575) | | | | 0.045 (*p* = .347) | | | | 0.222 (*p* < **.001**) | | | |

β = beta coefficient. CR = cognitive reserve. MoCA = Montreal Cognitive Assessment. MRI = magnetic resonance imaging. *p* = *p*-value. $R^2$ = proportion of variance explained by predictor. SE = standard error.

**Table 8**

Regressions for specific motor function tests (uncorrected p-values ≤ .05 are shown in bold).

| | Tapping test score | | | | Two-choice response time | | | | Tremor score (n=100) | | | | Balance score (n=100) | | | |
|---|---|---|---|---|---|---|---|---|---|---|---|---|---|---|---|---|
| | β | SE | *p* | $R^2$ | β | SE | *p* | $R^2$ | β | SE | *p* | $R^2$ | β | SE | *p* | $R^2$ |
| CR | | | | | | | | | | | | | | | | |
| CR proxy | 3.893 | 1.327 | **.004** | 0.083 | 10.58 | 4.312 | **.016** | 0.059 | 0.079 | 0.044 | .075 | 0.033 | 0.041 | 0.038 | .286 | 0.012 |
| MRI brain measures | | | | | | | | | | | | | | | | |
| Brain Structure | 0.712 | 0.402 | .080 | 0.027 | -0.502 | 1.306 | .702 | 0.001 | 0.005 | 0.013 | .682 | 0.001 | 0.002 | 0.011 | .882 | 0.000 |
| Brain Diffusion | -0.148 | 0.421 | .726 | 0.001 | 2.439 | 1.368 | .078 | 0.032 | -0.009 | 0.014 | .510 | 0.003 | -0.002 | 0.012 | .843 | 0.000 |
| Brain Functional | 0.865 | 0.508 | .091 | 0.023 | 1.134 | 1.649 | .493 | 0.006 | 0.026 | 0.017 | .126 | 0.023 | -0.007 | 0.014 | .654 | 0.002 |
| Total Model $R^2$ | 0.134 (*p* = **.008**) | | | | 0.097 (*p* = **.041**) | | | | 0.061 (*p* = .198) | | | | 0.015 (*p* = .836) | | | |

Two different subjects were identified as outliers for tremor and balance (exceeding three standard deviations from the mean) and were therefore excluded from comparisons involving these specific outcomes. β = beta coefficient. CR = cognitive reserve. MRI = magnetic resonance imaging. *p* = p-value. $R^2$ = proportion of variance explained by predictor. SE = standard error.
